# Association of SARS-CoV-2 Nucleocapsid Protein Mutations with Patient Demographic and Clinical Characteristics during the Delta and Omicron Waves

**DOI:** 10.1101/2023.02.26.23285573

**Authors:** Feda A. Alsuwairi, Asma Alsaleh, Madain S. Alsanea, Ahmed A. Al-Qahtani, Dalia Obeid, Reem S. Almaghrabi, Basma M. Alahideb, Maha A. AlAbdulkareem, Maysoon S. Mutabagani, Sahar I. Althawadi, Sara A. Altamimi, Abeer N. Alshukairi, Fatimah S. Alhamlan

## Abstract

SARS-CoV-2 genomic mutations outside the spike protein that may increase transmissibility and disease severity have not been well characterized. This study identified mutations in the nucleocapsid protein and their possible association with patient characteristics. We analyzed 695 samples from patients with confirmed COVID-19 in Saudi Arabia between April 1, 2021, and April 30, 2022. Nucleocapsid protein mutations were identified through whole genome sequencing. *Χ*2 tests and T tests assessed associations between mutations and patient characteristics. Logistic regression estimated risk of intensive care unit (ICU) admission or death. Of 60 mutations identified, R203K was most common followed by G204R, P13L, and E31del, R32del, and S33del. These mutations were associated with reduced risk of ICU admission. P13L, E31del, R32del, and S33del were also associated with reduced risk of death. By contrast, D63G, R203M, and D377Y were associated with increased risk of ICU admission. Most mutations were detected in the SR-rich region, which was associated with low risk of death. C-tail and central linker regions were associated with increased risk of ICU admission, whereas the N-arm region was associated with reduced ICU admission risk. Some SARS-CoV-2 nucleocapsid amino acid mutations may enhance viral infection and COVID-19 disease severity.

## 1. Introduction

Severe acute respiratory syndrome coronavirus 2 (SARS□CoV□2) is the etiological agent of coronavirus disease 19 (COVID-19). The virus emerged in Wuhan, China, in December 2019. It evolved and spread rapidly, leading to the COVID-19 pandemic [1]. The rapid evolution of SARS□CoV□2 highlights the importance of monitoring and tracing the emerging mutations in the genome.

SARS-CoV-2 is an RNA virus that tends to accumulate mutations in the genome. Various mutations have formed new virus variants [2]. Thus, several global efforts have been devoted to tracking changes in the genome. The World Health Organization (https://www.who.int/), Pango (https://cov-lineages.org/), Nextstrain (https://nextstrain.org/), and the Global Initiative on Sharing Avian Influenza Data (GISAID) (https://gisaid.org/) established nomenclature systems for naming and tracking SARS-CoV-2 genetic lineages [3,4]. The main identified variants and subvariants of the virus that resulted in pandemic waves and millions of deaths around the world included Alpha (B.1.1.7), Beta (B.1.351), Gamma (P.1), Delta (B.1.617.2) and Omicron (B.1.1.529, BA.1, BA.2, BA.4, and BA.5 lineages) [5]. The naming and tracking of SARS-CoV-2 genetics have been assisted by the deposits of SARS□CoV□2 genomic sequences in public genome sharing databases, such as GISAID and the GeneBank of the National Center for Biotechnology Information. As of November 2022, >13 million complete and high-coverage genomes of SARS-CoV-2 were deposited in GISAID and are globally accessible, of which >1,500 sequences are from Saudi Arabia.

The SARS-CoV-2 genome encodes 14 open reading frames, which express approximately 29 structural and nonstructural proteins. The spike glycoprotein, the membrane protein, the envelop protein, and the nucleocapsid (N) protein are the four essential structural proteins [6]. The spike protein is distributed on the surface of the virus and is responsible for attaching and binding to the human receptor for angiotensin-converting enzyme 2 [7]. Thus, the spike protein has been the target of the developed vaccines and therapeutics. In addition, many studies have focused on tracking emerged mutations in the spike protein and their impacts on virus transmissibility, disease pathogenesis, and immunogenicity. For example, the P681R mutation was shown to increase virus transmissibility, and the E484K mutation to affect antibody neutralization [8,9]. However, in addition to these studies focused on the spike protein, research should also be conducted to detect and assess mutations in other virus proteins, for example, the N protein.

The N protein plays numerous essential roles in the infection cycle of the virus. One role is viral RNA assembly and packaging into the riboneucleocapsid protein complex. The N protein also facilitates viral RNA replication and translation [10]. These important functions involve two conserved structural regions: (1) the N-terminal domain, representing the RNA binding domain (residues 44-174), and (2) the C-terminal domain, representing the dimerization domain (residues 255-364). These two domains are flanked by three intrinsically disordered regions: (1) the N-arm (residues 1-43), (2) a central linker region (residues 175-254), and (3) the C-tail (residues 365-419). The central linker region contains an SR-rich motif, enriched in serine and arginine residues. It connects the N-terminal domain and the C-terminal domain, with the N-arm and C-tail existing at the sides of the N-terminal and C-terminal domains [11–13]. Owing to the important roles of these regions in the assembly and synthesis of viral RNA, surveillance of SARS-CoV-2 should include tracking the evolution of mutations in these regions and the influences of these mutations on the characteristics of the virus. Here we report amino acid changes in the N protein and the association of these mutations with the demographic and clinical characteristics of SARS-CoV-2–positive patients visiting the King Faisal Specialist Hospital and Research Centre (KFSHRC), a tertiary-referral hospital located in Saudi Arabia and providing specialized care for, among other specialties, cancer, transplantation, and individuals who are immunocompromised.

## 2. Methods

### 2.1. Collection of patient samples and demographic and clinical data

Nasopharyngeal swab samples that were polymerase chain reaction (PCR)-confirmed positive for SARS-CoV-2 were deidentified, coded, and obtained in viral transport media from the Microbiology Section of the Pathology and Laboratory Medicine Department at KFSHRC in Riyadh, Madinah, and Jeddah. Samples were collected from 695 patients during the Delta and Omicron variant waves, from April 1, 2021, to April 30, 2022. The related clinical and demographic electronic health records were obtained from the Infection Control and Hospital Epidemiology Department at KFSHRC.

### 2.2. Whole Genome Sequencing

Unless otherwise specified, all equipment and kits for whole genome sequencing were purchased from Thermo Fisher Scientific, USA. Total viral RNA was extracted from 200 μL of viral transport media using the MagMAX™ Viral/Pathogen II Nucleic Acid Isolation Kit. Detection of viral RNA and estimation of viral load were performed using a TaqPath™ COVID-19 CE-IVD RT-PCR Kit that targets the N, spike, and open reading frame 1ab genes in SARS-CoV-2. Extracts that were positive by real-time PCR were converted to cDNA using an Invitrogen SuperScript™ IV VILO™ Master Mix kit. The assays were conducted according to the manufacturer’s instructions. The cDNA was used to prepare libraries with the Ion AmpliSeq™ SARS-CoV-2 Research Panel. The preparation of the libraries included (1) amplification of the targets, (2) partial digestion of amplicons, and (3) ligation of adapters to the amplicons. For each sample, two separate amplification reactions were conducted, with primer pool 1 used in one reaction and primer pool 2 used in the other. The amplification reactions were then combined. The amplified cDNA target was then partial digested. The products were ligated with unique barcode adapters using Ion Xpress™ Barcode Adapters 1-96 Kits. Each library was purified using 45 µL (i.e., 1.5× sample volume) of the Agencourt™ AMPure™ XP Reagent (Beckman Coulter, USA). Library preparation was performed according to the Ion AmpliSeq™ Library Kit Plus User Guide (MAN0006735) and following the Ion AmpliSeq™ RNA Libraries protocol. All reactions were performed using a VeritiTM 96-well Thermal Cycler. All barcoded libraries were quantified using an Ion Library TaqMan Quantitation Kit, normalized from original libraries using nuclease-free water to 28-33 pM for the Ion 520-530 chip or to 50 pM for the Ion 540 chip and then pooled in equal volumes based on selected Ion chip capacity, prior to undergoing automated template preparation. Template preparation included emulsion PCR and then immobilization of each DNA fragment on Ion Sphere™ Particles. These cloned DNA fragments were loaded into wells of an electronic semiconductor chip (Ion 520™ Chip, Ion 530™ Chip or Ion 540™ Chip). The automatic template preparation was performed using the Ion Chef™ Instrument with the Ion 510™ & Ion 520™ & Ion 530™ Kit or with the Ion 540™ Kit. Whole genome sequencing was performed with the Ion GeneStudio™ S5 System using Ion S5 sequencing solutions from the Ion 510™ & Ion 520™ & Ion 530™ Kit–Chef or the Ion 540™ Kit-Chef.

### 2.3. Data analysis

Torrent Suite™ Software version 5.12 (Thermo Fisher Scientific, USA) was used to analyze the sequencing data. The sequences were aligned with the Wuhan-Hu-1 reference genome (accession number MN908947.3). The de novo report of assembled contigs into FASTA file format was performed using the AssemblerTrinity plugin (v1.2.1.0). Metrics for quality control of each sequence were analyzed with the CoverageAnalysis plugin (v5.10.0.3). Single-nucleotide variants and indel variants were called using the variantCaller plugin (v5.10.1.19). Variant call files were analyzed with the COVID19AnnotateSnpEff plugin (v1.2.1.0) to predict the effects of mutations on the nucleic acid level and on the amino acid level. FASTA files of consensus sequences were generated using the IRMAreport plugin (v1.2.1.0). Consensus sequences were analyzed with the Nextclade web tool v2.8.1 (https://nextstrain.org) to align N gene sequences from our population and compare them with Wuhan-Hu-1 sequence.

### 2.4. Statistical analysis

The initial number of samples sequenced was 712. Samples were checked for duplicates, and patients with identical sequencing results were removed or merged. The final number of samples included in the study was 695. Data were cleaned and analyzed using SAS, version 9.4, and Prism, version 9.0 (GraphPad). Inferential and descriptive statistics were conducted to assess clinical variables. T tests were used to assess continuous variables, and χ2 tests were used to assess categorical variables. Logistic regression was conducted to estimate the risk of intensive care unit (ICU) admission and of death. All reported p values were two-tailed and were considered to be statistically significant at <0.05.

### 2.5. Data availability

The data and codes used in this study are available on request. The SARS-CoV-2 sequences were deposited on the GISAID website.

## 3. Results

### 3.1 Patient demographic characteristics

Samples were collected from 695 patients beginning April 1, 2021 and ending April 30, 2022. Most samples were collected in January 2022 (47.9%), followed by June 2021 (11.2%), and most patients visited the hospital located in Riyadh (75.3%), followed by Jeddah (13.1%), and then Madinah (11.7%). The mean (SD) age of the patients was 38.9 (18.5) years, with the youngest being 3 weeks and the eldest being 102 years. By gender, 53.5% of the patients were female, and 46.5% were male. The majority of the patients were Saudi nationals (72.3%) and were non-smokers (92.6%). By the end of our study, 80.7% of patients had recovered without the need for hospitalization, 5.8% died, and the remaining patients were either discharged after recovering or still recovering at the hospital. In total, 55.7% of patients had no comorbidity, whereas 44.3% presented with comorbidities, including 22.6% who were immunocompromised, 15.5% with diabetes, 24.8% with hypertension, and 7.1% (48) with organ transplantation. Most patients were symptomatic (81.6%), with only 13.7% being asymptomatic. Most patients presented with mild disease severity (83.4%), followed by Stage C (10.7%) and Stage D (5.8%) disease severity. Most patients did not require hospitalization (80.0%), but some (9.5%) required a short period of hospitalization and about the same number (10.5%) required longer hospitalization although for <20 days. Of our cohort, 13.6% were admitted to the ICU. The viral load detected in most patients was moderate (cycle threshold [Ct]: 20-30) (61.8%), with 27% of patients showing high viral load (Ct <20). Most patients were vaccinated (58.7%), 7.2% were unvaccinated, but the vaccination status in 34.1% was unknown. The vaccines most frequently received were from Pfizer (45.6%) followed by AstraZeneca (32.4%), with 15.0% of patients receiving vaccines from more than one company, and the rest were unknown (7.1%). Most patients received a second dose (39.5%), with 33.6% receiving only the first dose but 22.5% receiving a booster dose.

### 3.2. Variants detected from April 1, 2021, through April 30, 2022

The most frequently detected variants in our population were Omicron (BA.1) at 59% of the samples, followed by Delta at 26%, Beta at 3.6%, and Alpha at 3.4% **(Table 1)**. Most cases were from samples collected during the Omicron wave (67.4%), followed by Delta wave (32.6%) **(Figure 1)**. The number of patients infected with Delta increased such that it was the predominant variant in June 2021 and remained circulating at a high proportion from August to November 2021. The number of patients infected with the Delta variant then declined in December 2021 and was replaced by infections with the Omicron BA.1 subvariant, which became the predominant variant in January 2022 and remained high at the end of our study on April 30, 2022.

**Table 1.** SAR-CoV-2 variants detected in patient samples collected from April 1, 2021, through April 30, 2022.

| **Variant** | **Frequency** | **Percentage** |
| --- | --- | --- |
| **Omicron BA.1** | 414 | 59.6% |
| **Delta** | 179 | 25.8% |
| **Alpha** | 24 | 3.46% |
| **Omicron BA.2** | 24 | 3.46% |
| **Other** | 24 | 3.46% |
| **Beta** | 23 | 3.30% |
| **Delta Plus** | 3 | 0.43% |
| **Eta** | 2 | 0.30% |
| **Kappa** | 1 | 0.30% |
| **Omicron** | 1 | 0.30% |

**Figure 1.**
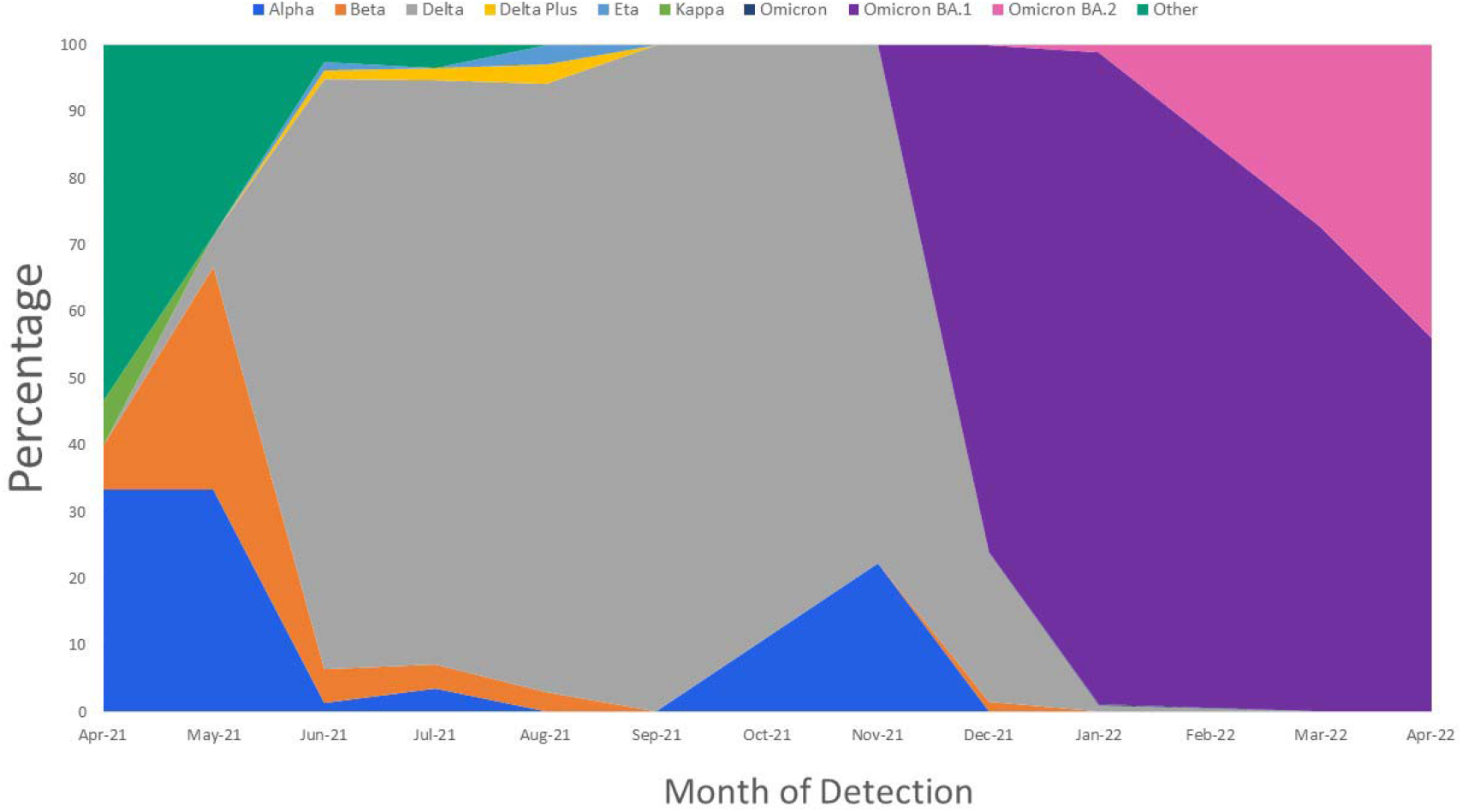
Wave distribution by month and variant detected.

### 3.3 Amino acid mutations detected in the N protein

Our analysis identified 60 amino acid mutations in the N protein derived from all patient samples. The most frequently mutated region within the protein was the SR-rich region, which represented 97.1% of the regions. The C-terminal domain had the fewest mutations (3.5%). A summary of the regions with amino acid mutations is given in **Table 2** and shown in **Figure 2**. All amino acids in the N protein that showed mutations are given in **Table 3** by frequency of mutation and percentage of the total sample. The most common amino acid mutations were R203K and G204R (both approximately 70%), followed by P13L, E31del, R32del, and S33del (all approximately 64%). **Figure 3** shows the frequency of the most common mutations detected by sample collection date across time. Overall, these mutations showed two similar patterns: the first included D377Y, D63G, G215C, and R203M, with detection frequencies peaking in June 2021 and then decreasing in January 2022 **(Figure 3A)**; the second pattern included R203K, E31del, R32del, S33del, G204R, and P13L, with detection frequencies peaking in January 2022 and remaining predominant at the study end (April 30, 2022) **(Figure 3B)**. The two consecutive mutations R203K and G204R were observed at the beginning of the study in April 2021, but then their frequency of detection declined in late June to re-emerged in December 2021 and peak in January 2022.

**Table 2.** Regions of the nucleocapsid (N) protein with amino acid mutations in this cohort.

| **N protein region** | **Frequency** | **Percentage** |
| --- | --- | --- |
| **N-arm**<br>(residues 1-43) | 471 | 67.8% |
| **SR-rich region**<br>(residues 175-204) | 675 | 97.1% |
| **NTD (RNA binding domain)**<br>(residues 44-174) | 216 | 31.1% |
| **LKR**<br>(residues 205-254) | 184 | 26.5% |
| **C-tail**<br>(residues 365-419) | 189 | 27.2% |
| **CTD (dimerization domain)**<br>(residues 255-364) | 24 | 3.5% |

Abbreviations: NTD, N-terminal domain; LKR, linker region; CTD, C-terminal domain.

**Table 3.** All detected amino acid mutations in the nucleocapsid protein by frequency and percentage of the total sample.

| **Amino Acid** | **Frequency** | **Percentage** |
| --- | --- | --- |
| **R203K** | 489 | 70.36% |
| **G204R** | 487 | 70.07% |
| **P13L** | 445 | 64.03% |
| **E31del** | 442 | 63.60% |
| **R32del** | 442 | 63.60% |
| **S33del** | 442 | 63.60% |
| **D63G** | 183 | 26.33% |
| **R203M** | 181 | 26.04% |
| **D377Y** | 179 | 25.76% |
| **G215C** | 132 | 18.99% |
| **S235F** | 24 | 3.45% |
| **T205I** | 24 | 3.45% |
| **D3L** | 23 | 3.31% |
| **I84V** | 22 | 3.17% |
| **T362I** | 12 | 1.73% |
| **A155V** | 4 | 0.58% |
| **K375N** | 4 | 0.58% |
| **A208V** | 3 | 0.43% |
| **D343G** | 3 | 0.43% |
| **P151L** | 3 | 0.43% |
| **Q9L** | 3 | 0.43% |
| **A12G** | 2 | 0.29% |
| **A134V** | 2 | 0.29% |
| **N354S** | 2 | 0.29% |
| **R385K** | 2 | 0.29% |
| **S105C** | 2 | 0.29% |
| **S202N** | 2 | 0.29% |
| **S2del+D3Y** | 2 | 0.29% |
| **T334R** | 2 | 0.29% |
| **A398S** | 1 | 0.14% |
| **D22Y** | 1 | 0.14% |
| **D348H** | 1 | 0.14% |
| **D399del** | 1 | 0.14% |
| **D3Y** | 1 | 0.14% |
| **G178C** | 1 | 0.14% |
| **G19V** | 1 | 0.14% |
| **G204L** | 1 | 0.14% |
| **G212C** | 1 | 0.14% |
| **G212V** | 1 | 0.14% |
| **G214+G215del** | 1 | 0.14% |
| **G238C** | 1 | 0.14% |
| **G30A** | 1 | 0.14% |
| **G71R** | 1 | 0.14% |
| **K361L** | 1 | 0.14% |
| **L113Q** | 1 | 0.14% |
| **N77Y** | 1 | 0.14% |
| **P279L** | 1 | 0.14% |
| **P279Q** | 1 | 0.14% |
| **P364L** | 1 | 0.14% |
| **Q244P** | 1 | 0.14% |
| **R32H** | 1 | 0.14% |
| **R40C** | 1 | 0.14% |
| **S201R** | 1 | 0.14% |
| **S327L** | 1 | 0.14% |
| **S79N** | 1 | 0.14% |
| **T141S** | 1 | 0.14% |
| **T247A** | 1 | 0.14% |
| **T366I** | 1 | 0.14% |
| **T379I** | 1 | 0.14% |
| **V72I** | 1 | 0.14% |

**Figure 2.**
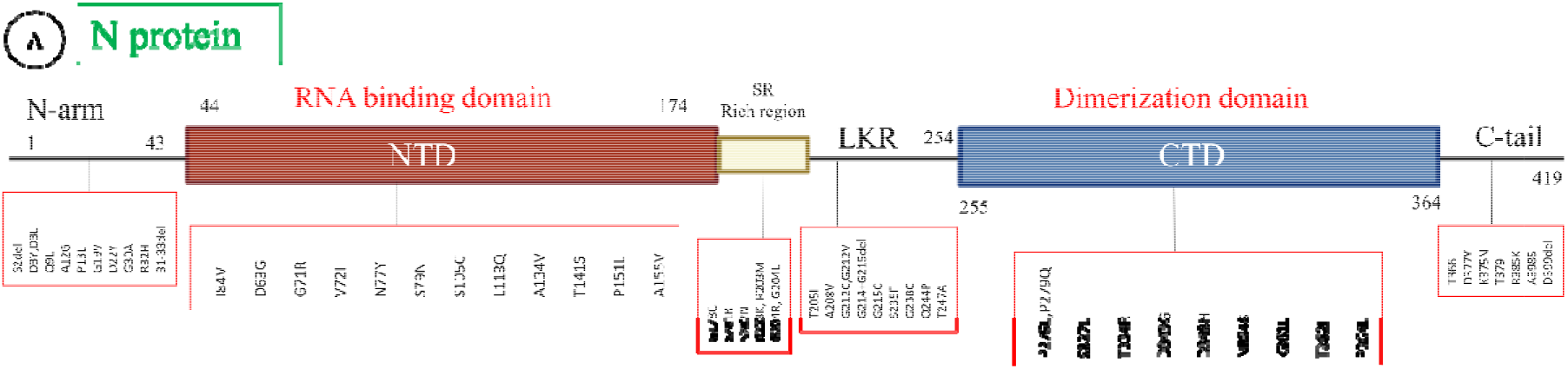
Distribution of the identified amino acid mutations across the various regions of the nucleocapsid (N) protein. NTD represents N-terminal domain; LKR, linker region; and CTD, C-terminal domain.

**Figure 3.**
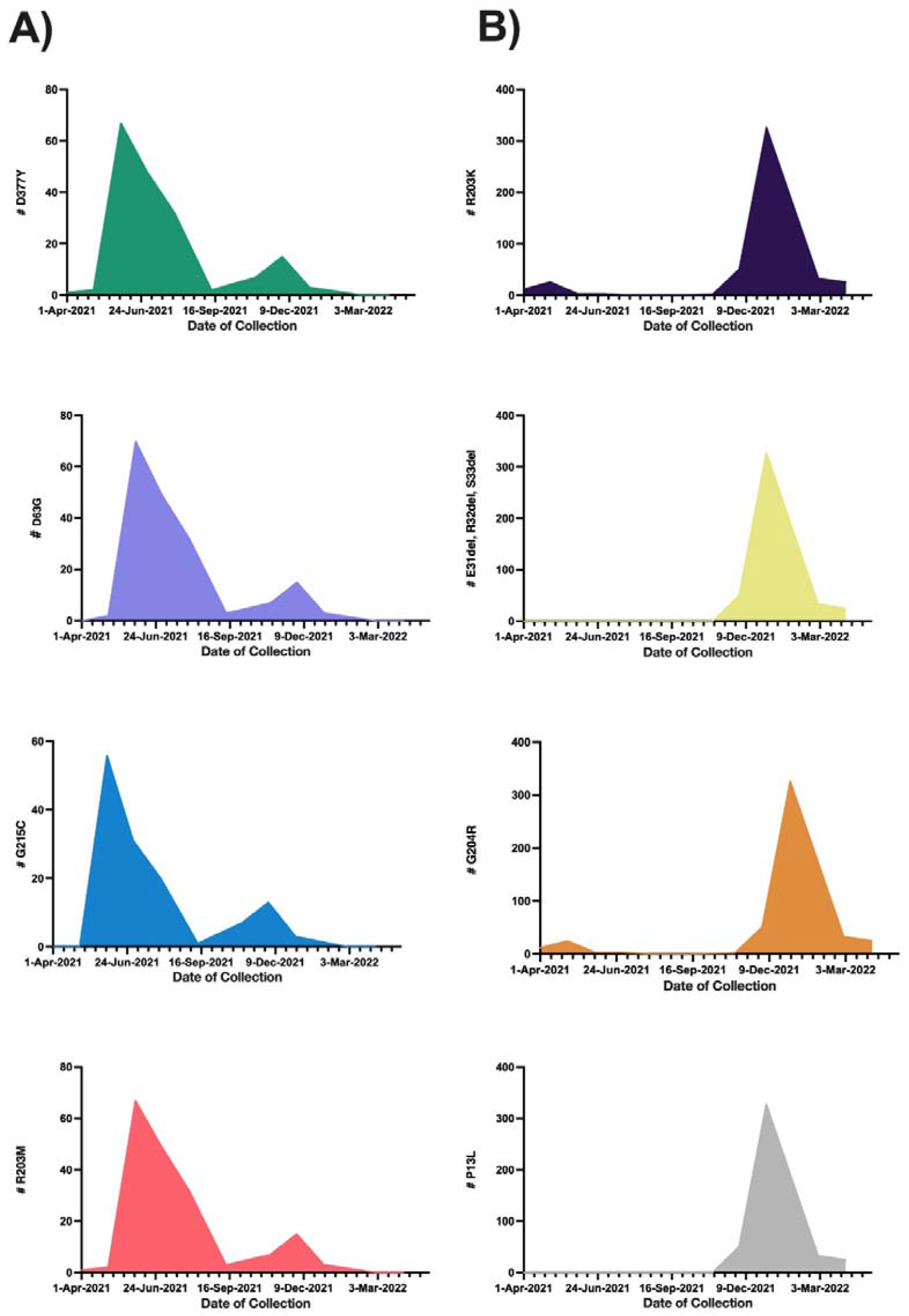
Distribution of the most frequently detected amino acid mutations in the nucleocapsid protein by date of sample collection. A) shows amino acid mutations with frequencies that peaked in June 2021 and decreased in April 2022. B) shows amino acid mutations with frequencies that peaked in January 2022.

### 3.4 Association of the most frequent amino acid mutations in the N protein with patient demographic and clinical characteristics

We used X2 tests and T tests to assess the associations between the most frequent amino acid mutations and patient demographic and clinical characteristics. Patients with the most common amino acid mutation (R203K) were significantly younger than patients with the wild type protein (P<0.05), and this mutation was detected more frequently in female patients than male patients (P<0.05) **(Table 4)**. The R203K mutation showed no association with rate of hospitalization or disease severity. No significant association was detected by vaccination status (i.e., vaccinated, unvaccinated, number of doses, and booster receipt). However, for the type of vaccine, most of the patients who received the Pfizer vaccine had the R203K mutation. By contrast, most patients with the wild type N protein and breakthrough infection received the AstraZeneca vaccine. Similar results were found for the G204R mutation. The results of the analyses for the associations of the remaining most frequently detected mutations are given in Supplemental **Tables 5-11**.

**Table 4.** Association of the nucleocapsid protein amino acid mutation R203K with patient demographic and clinical characteristics

| Characteristic | No.(%) | | $\chi^2$ or T (P-value) |
| --- | --- | --- | --- |
|  | R203K Mutation | Wild Type |  |
| **Age (mean, SD), years** | 36.6(18.7) | 44.3(16.8) | 5.3(<0.0001)* |
| **Variant** |  |  |  |
| Alpha | 24(3.5) | 0(0) | 672(<0.0001)* |
| Beta | 1(0.1) | 22(3.2) |  |
| Delta | 0(0) | 179(25.8) |  |
| Delta Plus | 0(0) | 3(0.4) |  |
| Eta | 0(0) | 2(0.3) |  |
| Kappa | 0(0) | 1(0.1) |  |
| Omicron | 0(0) | 1(0.1) |  |
| Omicron BA.1 | 412(59.3) | 2(0.3) |  |
| Omicron BA.2 | 24(3.5) | 0(0) |  |
| Other | 22(3.2) | 2(0.3) |  |
| **Wave** |  |  |  |
| Delta | 45(6.5) | 179(25.8) | 380(<0.001)* |
| Omicron | 438(63) | 33(4.5) |  |
| **Sex** |  |  |  |
| Male | 212(30.5) | 111(16.0) | 4.2(0.04)* |
| Female | 271(39.0) | 101(14.5) |  |
| **Nationality** |  |  |  |
| Saudi | 340(53.5) | 119(18.7) | 26.4(<0.001)* |
| Non-Saudi | 93(14.7) | 83(13.1) |  |
| Unknown=60 |  |  |  |
| **Smoking status** |  |  |  |
| Yes | 36(5.6) | 12(1.9) | 0.9(0.35) |
| No | 408(63.4) | 188(29.2) |  |
| Unknown=51 |  |  |  |
| **Patient Status** |  |  |  |
| Deceased | 24(3.5) | 16(2.3) | 9.3(0.026)* |
| Recovered | 398(57.8) | 158(22.9) |  |
| Hospitalized | 5(0.7) | 1(0.2) |  |
| Released | 50(7.3) | 37(5.4) |  |
| Unknown=6 |  |  |  |
| **Immunocompromised** |  |  |  |
| Yes | 358(53.5) | 45(6.7) | 0.065(0.80) |
| No | 106(23.9) | 160(23.9) |  |
| Unknown=26 |  |  |  |
| **ICU Admission** |  |  |  |
| Yes | 50(7.4) | 42(6.2) | 11.7(0.0006)* |
| No | 421(62.2) | 164(24.2) |  |
| Unknown=18 |  |  |  |
| **Comorbidity** |  |  |  |
| Yes | 213(31.8) | 84(12.5) | 1.3(0.25) |
| No | 252(37.6) | 121(18.1) |  |
| Unknown=25 |  |  |  |
| **Diabetes mellitus** |  |  |  |
| Yes | 59(8.8) | 45(6.7) | 9.5(0.002)* |
| No | 409(60.8) | 160(23.8) |  |
| Unknown=22 |  |  |  |
| **Hypertension** |  |  |  |
| Yes | 103(15.3) | 64(9.5) | 6.5(0.011)* |
| No | 365(54.2) | 141(21.0) |  |
| Unknown=22 |  |  |  |
| **Symptoms** |  |  |  |
| Asymptomatic | 30(5.0) | 2(0.3) | 10.8(0.001)* |
| Symptomatic | 374(62.2) | 195(32.5) |  |
| Unknown= 94 |  |  |  |
| **Disease Severity** |  |  |  |
| Mild | 374(59.0) | 155(24.5) | 16.4(0.0003)* |
| Stage C | 41(6.5) | 27(4.3) |  |
| Stage D | 15(2.4) | 22(3.5) |  |
| Unknown=61 |  |  |  |
| **Vaccination Status** |  |  |  |
| Vaccinated | 270(59.0) | 138(30.1) | 1.2(0.27) |
| Unvaccinated | 37(8.1) | 13(2.8) |  |
| Unknown= 237 |  |  |  |
| **Type of vaccine** |  |  |  |
| Pfizer | 141(37.2) | 45(11.9) | 81.6(<0.0001)* |
| AstraZeneca | 48(12.7) | 84(22.2) |  |
| Mixture | 58(15.3) | 3(0.8) |  |
| Unknown=316 |  |  |  |
| **Vaccine Dose** |  |  |  |
| Post first | 43(11.0) | 94(24.1) | 121.4(<0.0001)* |
| Post second | 125(32.1) | 36(9.2) |  |
| Post booster | 89(22.8) | 3(0.8) |  |
| Unknown=305 |  |  |  |
| **Hospitalization** |  |  |  |
| None | 396(57.9) | 151(22.1) | 10.1(0.0063)* |
| Short (≤20 days) | 38(5.6) | 27(4.0) | |
| Long ($>$20 days) | 42(6.1) | 30(4.4) | |
| Unknown=11 |  |  |  |
| **Organ Transplant** |  |  |  |
| Yes | 188(27.7) | 17(2.5) | 0.65(0.42) |
| No | 442(65.2) | 188(27.7) |  |
| Unknown=17 |  |  |  |
| **Ct Range** |  |  | 23.7(<0.0001)* |
| high Ct $>$30 | 53(8.13) | 20(3.07) | |
| Low Ct $<$20 | 96(14.72) | 80(12.27) | |
| Moderate Ct 20-30 | 301(46.17) | 102(15.64) |  |
| Unknown=43 |  |  |  |

Abbreviations: ICU, intensive care unit, Ct, cycle threshold. \*Significant P value, $P < 0.05$ .

### 3.5 Risk of ICU admission or death, by mutation

Logistic regression was used to estimate the risk of patient ICU admission or death **(Table 12)**. For ICU admission, lower risk was observed for females than males and for younger vs. older patients. Unvaccinated patients had higher risk of ICU admission (odds ratio=5.5, CI 95%: 2.7-10.8) than vaccinated patients. Patients with high viral load (low Ct) had higher risk of admission compared with patients who had moderate or low viral load. Patients with comorbidities, diabetes, or who were immunocompromised all showed a significant risk of ICU admission. The results of analyses by amino acid mutation indicated that patients with E31del, R32del, and S33del together or with R203K, G204R, and P13L, and were associated with reduced ICU admission risk compared with patients with the wild type N protein. Conversely, an increased risk for ICU admission was found for patients with amino acid mutations D63G, R203M, and D377Y. By N protein region, significant risk of ICU admission was found for patients with mutations located in the central linker region and the C-tail region, whereas patients with mutations located in the N-arm showed reduced risk.

**Table 12.** Estimated odds of ICU admission and death for patients with COVID-19, by patient characteristic and mutation.

| Characteristic | ICU Admission |  | Death |  |
| --- | --- | --- | --- | --- |
| | OR (95% CI) | $\chi^2$ (P-value) | OR (95% CI) | $\chi^2$ (P-value) |
| **Sex** |  |  |  |  |
| Male | 1 | 5.2(0.02) * | 1 | 7.6(0.0058)* |
| Female | 0.59(0.38-0.93) |  | 0.39(0.20-0.78) |  |
| **Age group, years** |  |  |  |  |
| 0-19 | 1 | 125.8(<0.001) * | 1 | 94.1(<0.001)* |
| 20-29 | 0.33(0.10-1.05) |  | NA |  |
| 30-39 | 0.23(0.067-0.78) |  | 0.24(0.02-2.6) |  |
| 40-49 | 1.22(0.48-3.1) |  | 1.32(0.24-7.4) |  |
| 50-59 | 1.44(0.56-3.73) |  | 1.9(0.34-10.7) |  |
| 60-102 | 10.8(4.624.9) |  | 18.8(4.3-81.8) |  |
| **Nationality** |  |  |  |  |
| Non-Saudi | 0.024(0.003-0.177) | 52.9(<0.0001) * | 0.067(0.009-0.49) | 17.4(<0.001)* |
| Saudi | 1 |  | 1 |  |
| **Vaccination status** |  |  |  |  |
| Vaccinated | 1 | 21.1(<0.0001) * | 1 | 3.3(0.068) |
| Unvaccinated | 5.5(2.7-10.81) |  | 2.9(1.0-8.4) |  |
| **Type of vaccine** |  |  |  |  |
| Pfizer | 5.4(1.58-18.61) | 12.0(0.0025) * | 6.7(0.83-53.2) | 5.5(0.063) |
| AstraZeneca | 1 |  | 1 |  |
| Mixture | 1.45(0.23-8.9) |  | 2.2(0.13-35.5) |  |
| **Vaccine dose** |  |  |  |  |
| Post first | 0.74(0.28-2.00) | 1.91(0.38) | 0.25(0.049-1.36) | 3.8(0.15) |
| Post second | 1.32(0.55-3.17) |  | 0.91(0.28-2.9) |  |
| Post booster | 1 |  | 1 |  |
| **Ct range** |  |  |  |  |
| high Ct >30 | 0.95(0.41-2.25) | 11.8(0.0027) * | 0.64(0.14-2.8) | 10.1(0.0065)* |
| Low Ct <20 | 2.36(1.4-3.8) |  | 2.7(1.4-5.4) |  |
| Moderate Ct 20-30 | 1 |  | 1 |  |
| **Hypertension** |  |  |  |  |
| No | 1 | 221.99(<0.0001) * | 1 | 30.9(<0.0001)* |
| Yes | 6.2(3.9-9.9) |  | 6.5(3.3-12.7) |  |
| **Diabetes mellitus** |  |  |  |  |
| No | 1 | 249.7(<0.0001) * | 1 | 50.0(<0.0001)* |
| Yes | 9.6(5.9-15.7) |  | 11.7(5.9-23.1) |  |
| **Comorbidity** |  |  |  |  |
| No | 1 | 146.8(<0.0001) * | NA | NA |
| Yes | 80.6(19.7-330.8) |  | NA |  |
| **Immunocompromised** |  |  |  |  |
| No | 1 | 179.9(<0.0001) * | 1 | 64.3(<0.0001) * |
| Yes | 30.95(17.1-55.9) |  | 17.1(7.7-38.2) |  |
| **Organ transplantation** |  |  |  |  |
| No | 1 | 132.9(<0.0001)* | 1 | 10.5(<0.001)* |
| Yes | 93.4(32.1-271.9) |  | 4.4(1.98-10.0) |  |
| **Amino acid mutation (Ref=0)** |  |  |  |  |
| **R203K** | 0.46(0.30-0.72) | 10.9(0.0009)* |  |  |
| **G204R** | 0.45(0.28-0.70) | 12.1(0.005)* | 0.58(0.30-1.1) | 2.6(0.10) |
| **P13L** | 0.37(0.230.58) | 19.1(<0.0001)* | 0.51(0.27-0.96) | 4.2(0.039)* |
| **E31del, R32del, S33del** | 0.37(0.24-0.59) | 18.8(<0.0001)* | 0.51(0.27-0.97) | 4.1(0.0417)* |
| **D63G** | 1.83(1.15-2.91) | 6.24(0.0125)* | 1.23(0.61-2.5) | 0.33(0.56) |
| **R203M** | 1.86(1.2-2.96) | 6.6(0.01)* | 0.64(0.33-1.23) | 1.72(0.18) |
| **D377Y** | 1.89(1.19-3.0) | 7.0(0.0081)* | 1.3(0.63-2.6) | 0.44(0.50) |
| **G215C** | 0.61(0.37-1.0) | 3.52(0.068) | 0.93(0.42-2.1) | 0.036(0.84) |
| **Mutation region (Ref=0)** |  |  |  |  |
| N-arm | 0.41(0.26-0.65) | 14.9(0.0001)* | 0.62(0.33-1.19) | 1.96(0.16) |
| SR-rich region | 0.35(0.13-0.04) | 3.7(0.053) | 0.23(0.072-0.71) | 4.9(0.026)* |
| NTD (RNA binding domain) | 2.1(1.3-3.3) | 10.1(0.0015)* | 1.7(0.9-3.2) | 2.5(0.11) |
| LKR | 2.2(1.4-3.5) | 10.95(0.0009)* | 1.54(0.78-3.0) | 1.5(0.22) |
| C-tail | 1.9(1.2-3.0) | 7.2(0.007)* | 1.2(0.57-2.3) | 0.16(0.68) |
| CTD (dimerization domain) | 1.7(0.62-4.7) | 0.98(0.32) | 1.5(0.34-6.7) | 0.27(0.61) |

Abbreviations: Ct, cycle threshold; CTD, C-terminal domain; ICU, intensive care unit; LKR, linker region; OR odds ratio; NTD, N-terminal domain; Ref, reference.

Regarding patient death, lower risk was observed for females vs. males and for younger vs. older patients. No significant risk of death was detected by vaccine status. Patients with a high viral load (low Ct) showed higher risk of death than patients with moderate or low viral load. Patients with comorbidities or diabetes or who were immunocompromised had a significant risk of death. The results of our analyses by mutation indicated that only patients with P13L or E31del, R32del, and S33del together were associated with reduced death risk compared with patients with the wild type N protein. By N protein region, significantly lower risk of death was found for patients with mutations located in the SR-rich region compared the wild type N protein.

## 4. Discussion

This epidemiologic surveillance study assessed the SARS-CoV-2 variants that were circulating at KFSHRC in Saudi Arabia and the evolution of N protein mutations and their association with patient characteristics and clinical data during the Delta and Omicron waves from April 1, 2021, to April 30, 2022. Our patient cohort showed increased frequency in the detection of the Delta (B.1.617.2) and Omicron (BA.1) variants from April 1, 2021, to April 31, 2022. Delta was the predominant variant in June 2021 but was replaced by Omicron in December 2021. These findings are in agreement with reported transmission rates from other regions, such as the United States, California, Germany, and Norway, with increases in the proportion of COVID-19 cases in December 2021 and with Omicron BA.1 being the dominant circulating variant. Thus, the findings of studies in other regions and in our findings in Saudi Arabia indicate that Omicron BA.1 has higher transmissibility than Delta [14–17].

We analyzed 695 SARS-CoV-2 sequences and identified 60 mutations in the N protein of this virus. The region in the protein with the most detected mutations was the SR-rich region, and the C-terminal domain had the fewest detected mutations. The SR-rich region is an intrinsically disordered region, and most of the protein’s phosphorylation sites are located in this region. Phosphorylation is important for the function of the N protein during RNA transcription [18,19]. Thus, a mutation in this region could alter phosphorylation. Previous studies have reported numerous important phosphorylation sites in the SR-rich region, including S184, T198, S201, S202, R203, and G204 [18,20]. In the present study, we detected six mutations in four of these phosphorylation sites: S201R, S202N, R203K, R203M, G204R and G204L. The R203K mutation was most frequently detected followed by the G204R mutation.

Both of them were observed in the Alpha variant and re-emerged with the Omicron variant. A similar observation was reported in other countries, including the United States [21]. Following these two mutations in predominance were P13L, and the E31del, R32del, S33del mutations, which peaked in frequency in January 2022. These mutations have been reported in other geographic regions [22,23]. P13 is located in an important T-cell epitope. Hence, a mutation in this position may change the properties of the epitope and the cellular immune response against the virus [24]. The P13L mutation and the E31del, R32del, and S33del mutations are located in the N-arm region, which is reported to function in regulating RNA binding; thus, these mutations may affect the regulation of RNA binding [25]. We observed a group of mutations (D377Y, D63G, G215C, and R203M) that peaked in June 2021 and decreased in December 2021. A similar observation has been reported for the same time frame in other countries, including the United States, Iran, Indonesia, and China [26–29].

We also observed mutation sets that were associated with specific patient characteristics. For example, the R203K, G204R, 31-33 del, and P13L mutations were detected in younger and female patients. These mutations were harbored in Omicron variants that observed in several studies to be associated with female and young ages [16,30,31].

In our cohort, patients who were unvaccinated, had a high viral load (low Ct), a comorbidity, or were immunocompromised showed significantly higher risk of ICU admission or death. There was no significant risk of death detected by vaccine status. A study conducted when Delta and Omicron dominated showed increased disease severity among unvaccinated individuals or persons with one or more comorbidity [32]. Other studies have found increased protection from virus infection or severe illness among individuals who received a booster and increased risk of COVID-19 disease among persons who are immunocompromised although this risk was reduced with vaccine receipt [33,34]. The association between high viral load (low Ct) and severe COVID□19 disease remains controversial. Consistent with our results, a study conducted in Japan examining the associations between viral load, COVID-19 disease severity, and clinical observations found that viral load was significantly higher in patients with severe disease or death than in patients with mild symptoms or asymptomatic cases [35]. By contrast, a study assessing randomly selected COVID-19 cases reported no association between disease severity and viral load although Ct values were lower for patients who were admitted without the need for supplemental oxygen than for patients who required oxygen support [36].

our assessments of the association of detected mutations with ICU admission and death risk indicated that D63G, R203M, and D377Y mutations were associated with increased risk for ICU admission. These mutations were reported in a case of breakthrough reinfection in which the patient experienced hypoxia and hospitalization, and the mutations were significantly associated with mortality [37]. These mutations may also play a role in spreading the virus. D63G was associated with a high viral load and may enhance virus immune escape during virus replication [38]. The mutation is located in the N-terminal domain of the N protein. The N-terminal domain interacts directly with RNA to form the ribonucleocapsid protein complex. Therefore, mutations in this region may impact the life cycle of the virus [39]. R203M and D377Y may affect the binding of antibodies [40]. In our study, R203K and G204R were associated with reduced risk of ICU admission, consistent with a similar observation by one study [41]. However, another study conducted in the U.S. showed an association of these mutations with severe disease and hospitalization [42]. In addition, these mutations were associated with higher viral loads among patients with COVID-19 in Saudi Arabia [43]. Our study also showed an association of P13L or of E31del, R32del, and S33del together with reduced risk of ICU admission or death compared with patients having the wild type N protein. These mutations have been detected in Hong Kong and were associated with mild COVID-19 disease in two cases [44].

The present study assessed the association of each region in the N protein showing mutations with risk of ICU admission or death. Among these regions, mutations in the C-tail and the central linker regions were significantly associated with ICU admission risk. The C-tail is located within the intrinsically disordered region of the N protein. The region is connected with C-terminal domain and thus may impact C-terminal domain oligomerization [13]. We detected seven mutations in the C-tail region. Among them, D377Y was the most frequent and was associated with ICU admission risk. The central linker region contains phosphorylation sites that are critical for regulating virus transcription and replication [19]. We detected nine mutations in this region. Of them, the G215C mutation was not significantly associated with risk of ICU admission or death, whereas mutations located in the N-arm region, including P13L and the E31del, R32del, and S33del mutations, showed reduced risk of ICU admission. We found that the SR-rich region was significantly associated with low risk of death. Although the R203M mutation in this region was associated with increased risk for ICU admission, two other mutations in the region, R203M and R203K, were associated with reduced risk of ICU admission.

### Limitations of the study

This study identified mutations located throughout the N protein and assessed the associations of these mutations with patient demographic and clinical characteristics. However, some detected mutations with no substantial expansion during the surveillance period were not included in our analyses. We focused on the patients’ vaccination state before the SARS-CoV-2 infection. However, the vaccine brand may not be associated with a specific mutation, which may have occurred in the variant before the vaccination. Information on patient condition, disease severity, vaccination status, and comorbidity, was not always complete and thus misclassification may have occurred, which may impact the results of the associations. The roles of some N protein regions are still unknown and the impact of the amino acid changes on the SARS-CoV-2 life cycle is ambiguous. Thus, future research on the structure and biochemistry of the protein is needed.

In conclusion, we reported amino acid changes in the N protein of SARS-CoV-2 that may impact virus transmission, infectivity, and immune escape. We found signification associations between both amino acid mutations and the regions of the N protein harboring the mutations with patient demographic and clinical characteristics. Additional monitoring of the evolution of the genetic changes in this SARS-CoV-2 protein may assist risk factor and therapeutic target identification as well as vaccine development and distribution.

## Data Availability

All data produced in the present study are available upon reasonable request to the authors

## Conflict of Interest

The authors declare that no conflicts of interest exist with regard to this study.

## Author Contributions

Conceptualization, F.A.A., M.S.A and F. S. A.; Methodology, F.A.A., A.N.A. and F. S. A.; Software, F.A.A. and B.M.A.; Validation, F.A.A., D.O., B.M.A. and M.A.A.; Formal Analysis, D.O.; Investigation, F.A.A., B.M.A., M.A.A., M.S.A and S.A.A; Resources, R.S. A., M.S.M., S.I.A. and A.N.A.; Data Curation, F.A.A., B.M.A., M.A.A. and M.S.A; Writing – Original Draft Preparation, F.A.A.; Writing – Review & Editing, F. S. A.; Visualization, F.A.A. and F. S. A.; Supervision, A.A.A. and F. S. A.; Project Administration, A.A.A. and F. S. A.; Funding Acquisition, A.A.A. and F. S. A. All authors have read and agreed to the published version of the manuscript.

## Funding

This work was supported by King Faisal Specialist Hospital and Research Centre [grant number 220 0009, 2020].

## Role of the Funder

The funder had no role in the study design; in the collection, analysis and interpretation of data; in the writing of the report; and in the decision to submit the article for publication.

## Ethical Approval

The study protocol was approved by the Ethics Committee of the Research Advisory Council at KFSHRC in Riyadh, Saudi Arabia (IRB No. 2200009), which also waived the requirement for obtaining patient consent because all data were deidentified.

## Notes

### Competing Interest Statement

The authors have declared no competing interest.

## References

1. Alnomasy, S. F. Virus-Receptor Interactions of SARS-CoV-2 Spikereceptor-Binding Domain and Human Neuropilin-1 B1 Domain. Saudi J. Biol. Sci. 2021, 28 (7), 3926–3928. https://doi.org/10.1016/j.sjbs.2021.03.074.

2. Cosar, B.; Karagulleoglu, Z. Y.; Unal, S.; Ince, A. T.; Uncuoglu, D. B.; Tuncer, G.; Kilinc, B. R.; Ozkan, Y. E.; Ozkoc, H. C.; Demir, I. N.; Eker, A.; Karagoz, F.; et al. SARS-CoV-2 Mutations and Their Viral Variants. Cytokine Growth Factor Rev. 2022, 63 (January), 10–22. https://doi.org/10.1016/j.cytogfr.2021.06.001.

3. Alm, E.; Broberg, E. K.; Connor, T.; Hodcroft, E. B.; Komissarov, Andrey B Maurer-Stroh, S.; Melidou, A.; Neher, R. A.; O’Toole, Á.; Pereyaslov, D. Geographical and Temporal Distribution of SARS-CoV-2 Clades in the WHO European Region, January to June 2020 Genome Data Availability during the COVID-19 Pandemic Clade Nomenclatures for SARS-CoV-2 Distribution of Clades over Time Clade Distribution in T. Eurosurveillance 2020, 25 (32), 2001410. https://doi.org/10.2807/1560-7917.ES.2020.25.32.2001410.

4. World Health Organization. Available online: https://www.who.int/ (accessed on 1 November 2022).

5. Centers for Disease Control and Prevention. Available online: https://www.cdc.gov/ (accessed on 1 November 2022).

6. Baruah, C.; Devi, P.; Sharma, D. K. Sequence Analysis and Structure Prediction of SARS-CoV-2 Accessory Proteins 9b and ORF14: Evolutionary Analysis Indicates Close Relatedness to Bat Coronavirus. Biomed Res. Int. 2020, 2020. https://doi.org/10.1155/2020/7234961.

7. Walls, A. C.; Park, Y.; Tortorici, M. A.; Wall, A.; Mcguire, A. T.; Veesler, D. Structure, Function, and Antigenicity of the SARS-CoV-2 Spike Glycoprotein. Cell 2020, 181 (2), 281–292.e6.

8. Cherian, S.; Potdar, V.; Jadhav, S.; Yadav, P.; Gupta, N.; Das, M.; Rakshit, P.; Singh, S.; Abraham, P.; Panda, S.; Team, N. I. C. SARS-CoV-2 Spike Mutations, L452R, T478K, E484Q and P681R, in the Second Wave of COVID-19 in Maharashtra, India. Microorganism 2021, 9 (7), 1–11. https://doi.org/https://doi.org/10.3390/microorganisms9071542.

9. Jangra, S.; Ye, C.; Rathnasinghe, R.; Stadlbauer, D.; Alshammary, H.; Amoako, A. A.; Awawda, M. H.; Beach, K. F.; Bermúdez-González, M. C.; Chernet, R. L.; Eaker, L. Q.; et al. SARS-CoV-2 Spike E484K Mutation Reduces Antibody Neutralisation. The Lancet Microbe 2021, 2 (7), e283–e284. https://doi.org/10.1016/S2666-5247(21)00068-9.

10. Kang, S.; Yang, M.; Hong, Z.; Zhang, L.; Huang, Z.; Chen, X.; He, S.; Zhou, Z.; Zhou, Z.; Chen, Q.; Yan, Y.; Zhang, C.; Shan, H.; Chen, S. Crystal Structure of SARS-CoV-2 Nucleocapsid Protein RNA Binding Domain Reveals Potential Unique Drug Targeting Sites. Acta Pharm. Sin. B 2020, 10 (7), 1228–1238. https://doi.org/10.1016/j.apsb.2020.04.009.

11. Dang, M.; Song, J. CTD of SARS-CoV-2 N Protein Is a Cryptic Domain for Binding ATP and Nucleic Acid That Interplay in Modulating Phase Separation. Protein Sci. 2022, 31 (2), 345–356. https://doi.org/10.1002/pro.4221.

12. Nikolakaki, E.; Giannakouros, T. SR/RS Motifs as Critical Determinants of Coronavirus Life Cycle. Front. Mol. Biosci. 2020, 7 (8), 1–6. https://doi.org/10.3389/fmolb.2020.00219.

13. Bai, Z.; Cao, Y.; Liu, W.; Li, J. The Sars-Cov-2 Nucleocapsid Protein and Its Role in Viral Structure, Biological Functions, and a Potential Target for Drug or Vaccine Mitigation. Viruses 2021, 13 (6). https://doi.org/10.3390/v13061115.

14. Lambrou, A. S.; Shirk, P.; Steele, M. K.; Paul, P.; Paden, C. R.; Cadwell, B. Genomic Surveillance for SARS-CoV-2 Variants: Predominance of the Delta (B.1.617.2) and Omicron (B.1.1.529) Variants — United States, June 2021– January 2022. CDC Morb. Mortal. Wkly. Rep. 2022, 71 (6), 206–211.

15. Modes, M. E.; Directo, M. P.; Melgar, M.; Johnson, L. R.; Yang, H.; Chaudhary, P.; Bartolini, S.; Kho, N.; Noble, P. W.; Isonaka, S.; Chen, P. Clinical Characteristics and Outcomes Among Adults Hospitalized with Laboratory-Confirmed SARS-CoV-2 Infection During Periods of B.1.617.2 (Delta) and B.1.1.529 (Omicron) Variant Predominance. CDC Morb. Mortal. Wkly. Rep. 2022, 71 (6), 217–223.

16. Sievers, C.; Zacher, B.; Ullrich, A.; Huska, M.; Fuchs, S.; Buda, S.; Haas, W.; Diercke, M.; an der Heiden, M.; Kröger, S. SARS-CoV-2 Omicron Variants BA.1 and BA.2 Both Show Similarly Reduced Disease Severity of COVID-19 Compared to Delta, Germany, 2021 to 2022. Eurosurveillance 2022, 27 (2). https://doi.org/10.2807/1560-7917.ES.2022.27.22.2200396.

17. Veneti, L.; Bøås, H.; Kristoffersen, A. B.; Stålcrantz, J.; Bragstad, K.; Hungnes, O.; Storm, M. L.; Aasand, N.; Rø, G.; Starrfelt, J.; et al. Reduced Risk of Hospitalisation among Reported COVID-19 Cases Infected with the SARS-CoV-2 Omicron BA.1 Variant Compared with the Delta Variant, Norway, December 2021 to January 2022. Eurosurveillance 2022, 27 (4), 1–8. https://doi.org/10.2807/1560-7917.ES.2022.27.4.2200077.

18. Wu, J.; Zhong, Y.; Liu, X.; Lu, X.; Zeng, W.; Wu, C.; Xing, F.; Cao, L.; Zheng, F.; Hou, P.; Peng, H.; Li, C.; Guo, D. A Novel Phosphorylation Site in SARS-CoV-2 Nucleocapsid Regulates Its RNA-Binding Capacity and Phase Separation in Host Cells. J. Mol. Cell Biol. 2022, 14 (2), 18–21. https://doi.org/10.1093/jmcb/mjac003.

19. Carlson, C. R.; Asfaha, J. B.; Ghent, C. M.; Howard, C. J.; Hartooni, N.; Safari, M.; Frankel, A. D.; Morgan, D. O. Phosphoregulation of Phase Separation by the SARS-CoV-2 N Protein Suggests a Biophysical Basis for Its Dual Functions. Mol. Cell 2020, 80 (6), 1092–1103.e4. https://doi.org/10.1016/j.molcel.2020.11.025.

20. Tung, H. Y. L.; Limtung, P. Mutations in the Phosphorylation Sites of SARS-CoV-2 Encoded Nucleocapsid Protein and Structure Model of Sequestration by Protein 14-3-3. Biochem. Biophys. Res. Commun. 2020, 532 (1), 134–138.

21. Johnson, B. A.; Zhou, Y.; Lokugamage, K. G.; Vu, M. N.; Bopp, N.; Crocquet-Valdes, P. A.; Kalveram, B.; Schindewolf, C.; Liu, Y.; Scharton, D.; et al. Nucleocapsid Mutations in SARS-CoV-2 Augment Replication and Pathogenesis. PLoS Pathog. 2022, 18 (6), 1–44. https://doi.org/10.1371/journal.ppat.1010627.

22. Chakraborty, C.; Bhattacharya, M.; Sharma, A. R.; Dhama, K.; Agoramoorthy, G. A Comprehensive Analysis of the Mutational Landscape of the Newly Emerging Omicron (B.1.1.529) Variant and Comparison of Mutations with VOCs and VOIs. GeroScience, 2022. https://doi.org/10.1007/s11357-022-00631-2.

23. Jung, C.; Kmiec, D.; Koepke, L.; Zech, F.; Jacob, T.; Sparrer, K. M. J.; Kirchhoff, F. Omicron: What Makes the Latest SARS-CoV-2 Variant of Concern So Concerning? J. Virol. 2022, 96 (6), 1–13. https://doi.org/10.1128/jvi.02077-21.

24. de Silva, T. I.; Liu, G.; Lindsey, B. B.; Dong, D.; Moore, S. C.; Hsu, N. S.; Shah, D.; Wellington, D.; Mentzer, A. J.; Angyal, A.; et al. The Impact of Viral Mutations on Recognition by SARS-CoV-2 Specific T Cells. iScience 2021, 24 (11). https://doi.org/10.1016/j.isci.2021.103353.

25. Hossain, A.; Akter, S.; Anjum, A.; Khair, S.; Rubayet, A. S. M. Unique Mutations in SARS-CoV-2 Omicron Subvariants’ Non-Spike Proteins: Potential Impacts on Viral Pathogenesis and Host Immune Evasion. Microb. Pathog. 2022, 170 (1). https://doi.org/doi:10.1016/j.micpath.2022.105699.

26. Kannan, S. R.; Spratt, A. N.; Cohen, A. R.; Naqvi, S. H.; Chand, H. S.; Quinn, T. P.; Lorson, C. L.; Byrareddy, S. N. Evolutionary Analysis of the Delta and Delta Plus Variants of the SARS-CoV-2 Viruses. J. Autoimmun. 2021, 124 (1).

27. Yavarian, J.; Nejati, A.; Salimi, V.; Jandaghi, N. Z. S.; Sadeghi, K.; Abedi, A.; Zarchi, A. S.; Gouya, M. M.; Mokhtari-Azad, T. Whole Genome Sequencing of SARS-CoV2 Strains Circulating in Iran during Five Waves of Pandemic. PLoS One 2022, 17 (5), 1–15. https://doi.org/10.1371/journal.pone.0267847.

28. Lie, E. L.; Hermawan, T. D. F.; Audah, K. A. Whole Genome Sequences Analyses of Indonesian Isolates SARS-CoV-2 Variants and Their Clinical Manifestations. Res. Sq. 2022, (submitted).

29. Feng, Z.; Cui, S.; Lyu, B.; Liang, Z.; Li, F.; Shen, L.; Xu, H.; Yang, P.; Wang, Q.; Zhang, D.; Pan, Y. Genomic Characteristics of SARS-CoV-2 in Beijing, China, 2021. Biosaf. Heal. 2022, 4 (4), 253–257. https://doi.org/10.1016/j.bsheal.2022.04.006.

30. Joung, S. Y.; Ebinger, J. E.; Sun, N.; Liu, Y.; Wu, M.; Tang, A. B.; Prostko, J. C.; Frias, E. C.; Stewart, J. L.; Sobhani, K.; Cheng, S. Awareness of SARS-CoV-2 Omicron Variant Infection among Adults with Recent COVID-19 Seropositivity. JAMA Netw. Open 2022, 5 (8), E2227241. https://doi.org/10.1001/jamanetworkopen.2022.27241.

31. Wolter, N.; Jassat, W.; Walaza, S.; Welch, R.; Moultrie, H.; Groome, M.; Amoako, D. G.; Everatt, J.; Bhiman, J. N.; Scheepers, C.; et al. Early Assessment of the Clinical Severity of the SARS-CoV-2 Omicron Variant in South Africa: A Data Linkage Study. Lancet 2022, 399 (10323), 437–446. https://doi.org/10.1016/S0140-6736(22)00017-4.

32. Kahn, F.; Bonander, C.; Moghaddassi, M.; Rasmussen, M.; Malmqvist, U.; Inghammar, M.; Bjork, J. Risk of Severe COVID-19 from the Delta and Omicron Variants in Relation to Vaccination Status, Sex, Age and Comorbidities - Surveillance Results from Southern Sweden, July 2021 to January 2022. Eurosurveillance 2022, 27 (9), 1–6. https://doi.org/10.2807/1560-7917.ES.2022.27.9.2200121.

33. Spitzer, A.; Angel, Y.; Marudi, O.; Zeltser, D.; Saiag, E.; Goldshmidt, H.; Goldiner, I.; Stark, M.; Halutz, O.; Gamzu, R.; Slobodkin, M.; et al. Association of a Third Dose of BNT162b2 Vaccine with Incidence of SARS-CoV-2 Infection among Health Care Workers in Israel. JAMA - J. Am. Med. Assoc. 2022, 327 (4), 341–349. https://doi.org/10.1001/jama.2021.23641.

34. Singson, J. R. C.; Kirley, P. D.; Pham, H.; Rothrock, G.; Armistead, I.; Meek, J.; Anderson, E. J.; Reeg, L.; Lynfield, R.; Ropp, S.; et al. Factors Associated with Severe Outcomes Among Immunocompromised Adults Hospitalized for COVID-19 — COVID-NET, 10 States, March 2020–February 2022. CDC Morb. Mortal. Wkly. Rep. 2022, 71 (27), 878–884. https://doi.org/10.15585/mmwr.mm7127a3.

35. Tsukagoshi, H.; Shinoda, D.; Saito, M.; Okayama, K.; Kimura, H.; Saruki, N. Relationships between Viral Load and the Clinical Course of COVID-19. Viruses 2021, 13 (2), 2–7. https://doi.org/https://doi.org/10.3390/v13020304.

36. Abdulrahman, A.; Mallah, S. I.; Alqahtani, M. COVID-19 Viral Load Not Associated with Disease Severity: Findings from a Retrospective Cohort Study. BMC Infect. Dis. 2021, 21 (1), 1–4. https://doi.org/10.1186/s12879-021-06376-

37. Shastri, J.; Parikh, S.; Aggarwal, V.; Agrawal, S.; Chatterjee, N.; Shah, R.; Devi, P.; Mehta, P.; Pandey, R. Severe SARS-CoV-2 Breakthrough Reinfection With Delta Variant After Recovery From Breakthrough Infection by Alpha Variant in a Fully Vaccinated Health Worker. Front. Med. 2021, 8 (8), 1–13. https://doi.org/10.3389/fmed.2021.737007.

38. Liu, W.; Li, H. COVID-19: Attacks Immune Cells and Interferences with Antigen Presentation through MHC-Like Decoy System. ChemRxiv 2021, (submitted).

39. Sarkar, S.; Runge, B.; Russell, R. W.; Movellan, K. T.; Calero, D.; Zeinalilathori, S.; Quinn, C. M.; Lu, M.; Calero, G.; Gronenborn, A. M.; Polenova, T. Atomic-Resolution Structure of SARS-CoV □ 2 Nucleocapsid Protein N □ Terminal Domain. J. Am. Chem. Soc. 2022, 144 (23). https://doi.org/10.1021/jacs.2c03320.

40. Moody, R.; Wilson, K. L.; Boer, J. C.; Holien, J. K.; Flanagan, K. L.; Jaworowski, A.; Plebanski, M. Predicted B Cell Epitopes Highlight the Potential for COVID-19 to Drive Self-Reactive Immunity. Front. Bioinforma. 2021, 1 (8), 1–21. https://doi.org/10.3389/fbinf.2021.709533.

41. Leary, S.; Gaudieri, S.; Parker, M. D.; Chopra, A.; James, I.; Pakala, S.; Alves, E.; John, M.; Lindsey, B. B.; Keeley, A. J.; et al. Generation of a Novel Sars-Cov-2 Sub-Genomic Rna Due to the R203k/G204r Variant in Nucleocapsid: Homologous Recombination Has Potential to Change Sars-Cov-2 at Both Protein and Rna Level. Pathog. Immun. 2021, 6 (2), 27–49. https://doi.org/10.20411/pai.v6i2.460.

42. Zhao, L. P.; Roychoudhury, P.; Gilbert, P.; Schiffer, J.; Lybrand, T. P.; Payne, T. H.; Randhawa, A.; Thiebaud, S.; Mills, M.; Greninger, A.; Pyo, C. W.; Wang, R.; Li, R.; Thomas, A.; Norris, B.; Nelson, W. C.; Jerome, K. R.; Geraghty, D. E. Mutations in Viral Nucleocapsid Protein and EndoRNase Are Discovered to Associate with COVID19 Hospitalization Risk. Sci. Rep. 2022, 12 (1), 1–11. https://doi.org/10.1038/s41598-021-04376-4.

43. Mourier, T.; Shuaib, M.; Hala, S.; Mfarrej, S.; Alofi, F.; Naeem, R.; Alsomali, A.; Jorgensen, D.; Subudhi, A. K.; Ben Rached, F.; Guan, Q.; et al, A. SARS-CoV-2 Genomes from Saudi Arabia Implicate Nucleocapsid Mutations in Host Response and Increased Viral Load. Nat. Commun. 2022, 13 (1). https://doi.org/10.1038/s41467-022-28287-8.

44. Gu, H.; Krishnan, P.; Ng, D. Y. M.; Chang, L. D. J.; Liu, G. Y. Z.; Cheng, S. S. M.; Hui, M. M. Y.; Fan, M. C. Y.; Wan, J. H. L.; Lau, L. H. K.; Cowling, B. J.; Peiris, M.; Poon, L. L. M. Probable Transmission of SARS-CoV-2 Omicron Variant in Quarantine Hotel, Hong Kong, China, November 2021. Emerg. Infect. Dis. 2022, 28 (2), 460–462. https://doi.org/10.3201/eid2802.212422.

