## Supplemental Tables 5-11 for "Association of SARS-CoV-2 Nucleocapsid Protein Mutations with Patient Demographic and Clinical Characteristics during the Delta and Omicron Waves"

Table 5. Association of the nucleocapsid protein amino acid mutation G204R with patient demographic and clinical characteristics

| **Characteristic** | **No.(%)** | | **χ^2^ or T** |
| --- | --- | --- | --- |
|  | **G204R Mutation** | **Wild Type** | **(P-value)** |
| **Age (mean, SD), years** | 36.6(18.7) | 44.3(16.8) | 5.4(<0.0001)* |
| **Variant** |  |  |  |
| Alpha | 24(3.5) | 0(0) | 665.5(<0.0001)* |
| Beta | 1(0.1) | 22(3.2) |  |
| Delta | 0(0.0) | 179(25.8) |  |
| Delta Plus | 0(0.0) | 3(0.4) |  |
| Eta | 0(0.0) | 2(0.3) |  |
| Kappa | 0(0.0) | 1(0.1) |  |
| Omicron | 0(0.0) | 1(0.1) |  |
| Omicron BA.1 | 412(59.3) | 2(0.3) |  |
| Omicron BA.2 | 24(3.5) | 0(0.0) |  |
| Other | 20(2.9) | 4(0.6) |  |
| **Wave** |  |  |  |
| Delta | 43(6.19) | 181(26.04) | 387(<0.0001)* |
| Omicron | 438(63.02) | 33(4.75) |  |
| **Sex** |  |  |  |
| Male | 211(38.9) | 112(14.7) | 3.9(0.047)* |
| Female | 270 (30.4) | 102(14.7) |  |
| **Nationality** |  |  |  |
| Saudi | 338(53.2) | 121(19.1) | 24.4 (<0.0001)* |
| Non-Saudi | 93(14.7) | 83(13.1) |  |
| Unknown=60 |  |  |  |
| **Smoking status** |  |  |  |
| Yes | 36(5.6) | 12(1.9) | 0.97(0.32) |
| No | 406(63.0) | 190(9.5) |  |
| Unknown=51 |  |  |  |
| **Patient Status** |  |  |  |
| Deceased | 17(2.47) | 23(3.34) | 9.9(0.02)* |
| Recovered | 159(23.08) | 397(57.62) |  |
| Hospitalized | 1(0.15) | 5(0.73) |  |
| Released | 37(5.37) | 50(7.26) |  |
| Unknown=6  UNK= |  |  |  |
| **Immunocompromised** |  |  |  |
| Yes | 105(15.7) | 46(6.9) | 0.02(0.90) |
| No | 357(53.4) | 161(24.1) |  |
| Unknown=26 |  |  |  |
| **ICU Admission** |  |  |  |
| Yes | 49(7.2) | 43(6.4) | 12.8(0.0003)* |
| No | 420(62.0) | 165(24.4) |  |
| Unknown=18 |  |  |  |
| **Comorbidity** |  |  |  |
| Yes | 122(31.5) | 85(12.7) | 1.3(0.26) |
| No | 251(37.5) | 122(18.2) |  |
| Unknown=25 |  |  |  |
| **Diabetes mellitus** |  |  |  |
| Yes | 58(8.6) | 46(6.8) | 10.5(0.0012)* |
| No | 408(60.6) | 161(23.9) |  |
| Unknown=22 |  |  |  |
| **Hypertension** |  |  |  |
| Yes | 102(15.2) | 46(9.7) | 6.9(0.0084)* |
| No | 364(54.1) | 161(21.1) |  |
| Unknown=22 |  |  |  |
| **Symptoms** |  |  |  |
| Asymptomatic | 30(5.0) | 2(0.3) | 11.0(0.0009)* |
| Symptomatic | 372(61.9) | 197(32.8) |  |
| Unknown=94 |  |  |  |
| **Disease Severity** |  |  |  |
| Mild | 373(58.8) | 156(24.6) | 18.6(<0.0001)* |
| Stage C | 41(6.5) | 27(4.3) |  |
| Stage D | 14(2.2) | 23(3.6) |  |
| Unknown=61 |  |  |  |
| **Vaccination Status** |  |  |  |
| Vaccinated | 270(59.0) | 138(30.1) | 0.29(0.6) |
| Unvaccinated | 138(7.6) | 15(3.3) |  |
| Unknown=237 |  |  |  |
| **Type of vaccine** |  |  |  |
| Pfizer | 141(37.2) | 45(11.9) | 81.6(<0.0001) * |
| AstraZeneca | 48(22.2) | 84(12.7) |  |
| Mixture | 58(15.3) | 3(0.80 |  |
| Unknown=316 |  |  |  |
| **Vaccine Dose** |  |  |  |
| Post first | 43(11.03) | 94(24.1) | 121.4(<0.0001)* |
| Post second | 125(32.05) | 36(9.23) |  |
| Post booster | 89(22.82) | 3(0.77) |  |
| Unknown=305 |  |  |  |
| **Hospitalization Duration** |  |  |  |
| None | 151(22.1) | 396(57.9) | 12.4(0.002)* |
| Short (≤20 days) | 38(5.9) | 27(4.7) |  |
| Long (>20 days) | 40(5.6) | 32(4.0) |  |
| Unknown=11 |  |  |  |
| **Organ Transplant patient** |  |  |  |
| Yes | 30(4.4) | 18(2.7) | 1.2(0.28) |
| No | 441(65.0) | 189(27.9) |  |
| Unknown=17 |  |  |  |
| **Ct Range** |  |  |  |
| high Ct >30 | 53(11.2) | 20(8.13) | 24.4(<0.0001)* |
| Low Ct <20 | 95(14.57) | 81(12.42) |  |
| Moderate Ct 20-30 | 300(46.01) | 103(15.8) |  |
| Unknown=43 |  |  |  |

Abbreviations: ICU, intensive care unit, Ct, cycle threshold. *Significant P value, P<0.05.

Table 6. Association of the nucleocapsid protein amino acid mutations E31del, R32del, and S33del with patient demographic and clinical characteristics

| **Characteristic** | **No.(%)** | | **χ^2^ or T** |
| --- | --- | --- | --- |
|  | **Del Mutation** | **Wild Type** | **(P-value)** |
| **Age (mean, SD), years** | 36.2(18.7) | 43.7(17.1) | 5.4(<0.0001)* |
| **Variant** |  |  |  |
| Alpha | 0(0) | 24(3.45) | 690.7(<0.0001)* |
| Beta | 0(0) | 23(3.31) |  |
| Delta | 0(0) | 179(25.76) |  |
| Delta Plus | 0(0) | 3(0.43) |  |
| Eta | 0(0) | 2(0.29) |  |
| Kappa | 0(0) | 1(0.14) |  |
| Omicron | 0(0) | 1(0.14) |  |
| Omicron BA.1 | 413(59.42) | 1(0.14) |  |
| Omicron BA.2 | 24(3.45) | 0(0) |  |
| Other | 0(0) | 24(3.45) |  |
| **Wave** |  |  |  |
| Delta | 0 | 224(32.2) | 559.9(<0.0001)* |
| Omicron | 437(62.9) | 34(4.9) |  |
| **Sex** |  |  |  |
| Male | 190(27.3) | 133(19.1) | 4.2(0.04)* |
| Female | 247(35.5) | 125(18.0) |  |
| **Nationality** |  |  |  |
| Saudi | 310(48.8) | 149(23.5) | 28.9 (<0.0001)* |
| Non-Saudi | 78(12.3) | 98(15.4) |  |
| Unknown=60 |  |  |  |
| **Smoking status** |  |  |  |
| Yes | 32(5.0) | 16(2.5) | 0.42(0.51) |
| No | 369(57.3) | 227(35.3) |  |
| Unknown=51 |  |  |  |
| **Patient Status** |  |  |  |
| Deceased | 19(2.76) | 21(3.05) | 8.9(0.03)* |
| Recovered | 362(52.54) | 194(28.16) |  |
| Hospitalized | 4(0.58) | 2(0.29) |  |
| Released | 46(6.68) | 41(5.95) |  |
| Unknown=6  Unknown= |  |  |  |
| **Immunocompromised** |  |  |  |
| Yes | 93(13.9) | 58(8.7) | 0.11(0.73) |
| No | 327(48.9) | 191(28.6) |  |
| Unknown=26 |  |  |  |
| **ICU Admission** |  |  |  |
| Yes | 39(5.8) | 53(7.8) | 19.5(<0.0001)* |
| No | 388(57.3) | 197(29.1) |  |
| Unknown=18 |  |  |  |
| **Comorbidity** |  |  |  |
| Yes | 188(28.1) | 109(16.3) | 0.05(0.82) |
| No | 233(34.8) | 140(20.9) |  |
| Unknown=25 |  |  |  |
| **Diabetes mellitus** |  |  |  |
| Yes | 51(7.6) | 53(7.9) | 10.3(0.0013)* |
| No | 373(55.2) | 196(29.1) |  |
| Unknown=22 |  |  |  |
| **Hypertension** |  |  |  |
| Yes | 92(13.7) | 75(11.1) | 5.9(0.015)* |
| No | 332(49.3) | 174(25.9) |  |
| Unknown=22 |  |  |  |
| **Symptoms** |  |  |  |
| Asymptomatic | 25(4.2) | 7(1.2) | 4.5(0.034)* |
| Symptomatic | 337(56.1) | 232(38.6) |  |
| Unknown= 94 |  |  |  |
| **Disease Severity** |  |  |  |
| Mild | 342(53.9) | 187(29.5) | 21.6(<0.0001)* |
| Stage C | 34(5.4) | 34(5.4) |  |
| Stage D | 11(1.7) | 26(4.1) |  |
| Unknown=61 |  |  |  |
| **Vaccination Status** |  |  |  |
| Vaccinated | 250(54.6) | 158(34.5) | 1.6(0.21) |
| Unvaccinated | 26(5.7) | 24(5.2) |  |
| Unknown= 237 |  |  |  |
| **Type of vaccine** |  |  |  |
| Pfizer | 135(35.6) | 51(13.5) | 111.2(<0.0001) * |
| AstraZeneca | 34(9.0) | 98(25.9) |  |
| Mixture | 59(15.6) | 2(0.5) |  |
| Unknown=316 |  |  |  |
| **Vaccine Dose** |  |  |  |
| Post first | 23(5.9) | 114(29.23) | 182.2(<0.0001)* |
| Post second | 126(32.31) | 35(8.97) |  |
| Post booster | 89(22.82) | 3(0.77) |  |
| Unknown=305 |  |  |  |
| **Hospitalization Duration** |  |  |  |
| None | 364(53.2) | 183(26.8) | 15.9(0.0003)* |
| Short (≤20 days) | 35(5.1) | 30(4.4) |  |
| Long (>20 days) | 32(4.7) | 40(5.9) |  |
| Unknown=11 |  |  |  |
| **Organ Transplant patient** |  |  |  |
| Yes | 26(3.8) | 22(3.2) | 1.8(0.17) |
| No | 403(59.4) | 227(33.5) |  |
| Unknown=17 |  |  |  |
| **Ct Range** |  |  |  |
| high Ct >30 | 45(6.9) | 28(4.3) | 18.1(0.0001)* |
| Low Ct <20 | 86(13.2) | 90(13.8) |  |
| Moderate Ct 20-30 | 272(41.7) | 131(20.1) |  |
| Unknown=43 |  |  |  |

Abbreviations: ICU, intensive care unit, Ct, cycle threshold. *Significant P value, P<0.05.

Table 7. Association of the nucleocapsid protein amino acid mutation P13L with patient demographic and clinical characteristics

| **Characteristic** | **No.(%)** | | **χ^2^ or T** |
| --- | --- | --- | --- |
|  | **P13L Mutation** | **Wild Type** | **(P-value)** |
| **Age (mean, SD), years** | 36.1(18.6) | 43.7(17.1) | 5.5(<0.0001)* |
| **Variant** |  |  |  |
| Alpha | 0(0) | 24(3.45) | 690.7(<0.0001)* |
| Beta | 0(0) | 23(3.31) |  |
| Delta | 0(0) | 179(25.76) |  |
| Delta Plus | 0(0) | 3(0.43) |  |
| Eta | 0(0) | 2(0.29) |  |
| Kappa | 0(0) | 1(0.14) |  |
| Omicron | 1(0.14) | 0(0) |  |
| Omicron BA.1 | 413(59.42) | 1(0.14) |  |
| Omicron BA.2 | 24(3.45) | 0(0) |  |
| Other | 0(0) | 24(3.45) |  |
| **Wave** |  |  |  |
| Delta | 0 | 224(32.2) | 563.3(<0.0001)* |
| Omicron | 438(63.0) | 33(4.5) |  |
| **Sex** |  |  |  |
| Male | 190(27.3) | 133(19.1) | 4.6(0.032)* |
| Female | 248(35.7) | 124(17.8) |  |
| **Nationality** |  |  |  |
| Saudi | 311(49.0) | 148(23.3) | 29.4 (<0.0001)* |
| Non-Saudi | 78(12.3) | 98(15.4) |  |
| Unknown=60 |  |  |  |
| **Smoking status** |  |  |  |
| Yes | 32(5.0) | 16(2.5) | 0.39(0.52) |
| No | 370(57.5) | 226(35.1) |  |
| Unknown=51 |  |  |  |
| **Patient Status** |  |  |  |
| Deceased | 19(2.76) | 21(3.05) | 9.2(0.02)* |
| Recovered | 363(52.7) | 193(28.0) |  |
| Hospitalized | 4(0.58) | 2(0.29) |  |
| Released | 46(6.68) | 41(5.95) |  |
| Unknown=6  Unknown= |  |  |  |
| **Immunocompromised** |  |  |  |
| Yes | 93(13.9) | 58(8.7) | 0.15(0.70) |
| No | 328(49.0) | 190(28.4) |  |
| Unknown=26 |  |  |  |
| **ICU Admission** |  |  |  |
| Yes | 39(5.8) | 53(7.8) | 19.8(<0.0001)* |
| No | 389(57.5) | 196(29.0) |  |
| Unknown=18 |  |  |  |
| **Comorbidity** |  |  |  |
| Yes | 189(28.2) | 108(16.1) | 0.09(0.75) |
| No | 233(34.8) | 140(20.9) |  |
| Unknown=25 |  |  |  |
| **Diabetes mellitus** |  |  |  |
| Yes | 51(7.6) | 53(7.9) | 10.5(0.0012)* |
| No | 374(55.6) | 195(29.0) |  |
| Unknown=22 |  |  |  |
| **Hypertension** |  |  |  |
| Yes | 92(13.7) | 75(11.1) | 6.2(0.012)* |
| No | 333(49.5) | 173(25.7) |  |
| Unknown=22 |  |  |  |
| **Symptoms** |  |  |  |
| Asymptomatic | 25(4.2) | 7(1.2) | 4.5(0.034)* |
| Symptomatic | 337(56.1) | 232(38.6) |  |
| Unknown= 94 |  |  |  |
| **Disease Severity** |  |  |  |
| Mild | 343(54.1) | 186(29.3) | 21.9(<0.0001)* |
| Stage C | 34(5.4) | 34(5.4) |  |
| Stage D | 11(1.7) | 26(4.1) |  |
| Unknown=61 |  |  |  |
| **Vaccination Status** |  |  |  |
| Vaccinated | 252(55.0) | 156(34.0) | 1.8(0.18) |
| Unvaccinated | 26(5.7) | 24(5.2) |  |
| Unknown= 237 |  |  |  |
| **Type of vaccine** |  |  |  |
| Pfizer | 136(35.9) | 50(13.2) | 112.5(<0.0001) * |
| AstraZeneca | 34(9.0) | 98(25.9) |  |
| Mixture | 59(15.6) | 2(0.5) |  |
| Unknown=316 |  |  |  |
| **Vaccine Dose** |  |  |  |
| Post first | 23(5.9) | 114(29.2) | 185.6(<0.0001)* |
| Post second | 126(32.3) | 35(9.0) |  |
| Post booster | 90(23.1) | 2(0.5) |  |
| Unknown=305 |  |  |  |
| **Hospitalization Duration** |  |  |  |
| None | 365(53.4) | 182(26.6) | 16.2(0.0003)* |
| Short (≤20 days) | 35(5.1) | 30(4.4) |  |
| Long (>20 days) | 32(4.7) | 40(5.9) |  |
| Unknown=11 |  |  |  |
| **Organ Transplant patient** |  |  |  |
| Yes | 26(3.8) | 22(3.2) | 1.9(0.17) |
| No | 404(59.6) | 226(33.3) |  |
| Unknown=17 |  |  |  |
| **Ct Range** |  |  |  |
| high Ct >30 | 46(7.1) | 27(4.1) | 19.2(0.0001)* |
| Low Ct <20 | 85(13.0) | 91(14.0) |  |
| Moderate Ct 20-30 | 272(41.7) | 131(20.1) |  |
| Unknown=43 |  |  |  |

Abbreviations: ICU, intensive care unit, Ct, cycle threshold. *Significant P value, P<0.05.

Table 8. Association of the nucleocapsid protein amino acid mutation D63G with patient demographic and clinical characteristics

| **Characteristic** | **No.(%)** | | **χ^2^ or T** |
| --- | --- | --- | --- |
|  | **D63G Mutation** | **Wild Type** | **(P-value)** |
| **Age (mean, SD), years** | 43.9(16.1) | 37.2(18.9) | 4.6(<0.0001)* |
| **Variant** |  |  |  |
| Alpha | 0.0(0.0) | 24.0(3.5) | 689.8(<0.0001)* |
| Beta | 0.0(0.0) | 23.0(3.3) |  |
| Delta | 178.0(25.6) | 1.0(0.1) |  |
| Delta Plus | 3.0(0.4) | 0.0(0.0) |  |
| Eta | 0.0(0.0) | 2.0(0.3) |  |
| Kappa | 0.0(0.0) | 1.0(0.1) |  |
| Omicron | 0.0(0.0) | 1.0(0.1) |  |
| Omicron BA.1 | 0.0(0.0) | 414.0(59.6) |  |
| Omicron BA.2 | 0.0(0.0) | 24.0(3.5) |  |
| Other | 0.0(0.0) | 24.0(3.5) |  |
| **Wave** |  |  |  |
| Delta | 152(21.87) | 72(10.36) | 300.0(<0.0001)* |
| Omicron | 29(4.17) | 442(63.6) |  |
| **Sex** |  |  |  |
| Male | 96(13.8) | 227(32.7) | 4.2(0.04)* |
| Female | 85(12.2) | 287(41.3) |  |
| **Nationality** |  |  |  |
| Saudi | 99(15.6) | 360(56.7) | 25.5 (<0.0001)* |
| Non-Saudi | 73(11.5) | 103(16.2) |  |
| Unknown=60 |  |  |  |
| **Smoking status** |  |  |  |
| Yes | 8(1.2) | 40(6.2) | 2.5(0.11) |
| No | 162(25.2) | 434(67.4) |  |
| Unknown=51 |  |  |  |
| **Patient Status** |  |  |  |
| Deceased | 12(1.7) | 28(4.1) | 9.3(0.029)* |
| Recovered | 134(19.5) | 422(61.3) |  |
| Hospitalized | 1(0.2) | 5(0.7) |  |
| Released | 34(4.9) | 53(7.7) |  |
| Unknown=6  Unknown= |  |  |  |
| **Immunocompromised** |  |  |  |
| Yes | 37(5.5) | 114(17.0) | 0.27 (0.59) |
| No | 138(20.6) | 380(56.8) |  |
| Unknown=26 |  |  |  |
| **ICU Admission** |  |  |  |
| Yes | 34(5.0) | 58(8.6) | 6.6(0.01)* |
| No | 142(21.0) | 443(65.4) |  |
| Unknown=18 |  |  |  |
| **Comorbidity** |  |  |  |
| Yes | 70(10.5) | 227(33.9) | 1.8(0.20) |
| No | 105(15.7) | 268(40) |  |
| Unknown=25 |  |  |  |
| **Diabetes mellitus** |  |  |  |
| Yes | 36(5.4) | 68(10.1) | 4.7(0.03)* |
| No | 139(20.6) | 430(63.9) |  |
| Unknown=22 |  |  |  |
| **Hypertension** |  |  |  |
| Yes | 51(7.6) | 116(17.2) | 2.4(0.12) |
| No | 124(18.4) | 382(56.8) |  |
| Unknown=22 |  |  |  |
| **Symptoms** |  |  |  |
| Asymptomatic | 2(0.3) | 30(5.0) | 7.9(0.00049)* |
| Symptomatic | 166(27.6) | 403(67.1) |  |
| Unknown=94 |  |  |  |
| **Disease Severity** |  |  |  |
| Mild | 134(21.1) | 395(62.3) | 12.6(0.0019)* |
| Stage C | 22(3.5) | 46(7.3) |  |
| Stage D | 19(3.0) | 18(2.8) |  |
| Unknown=61 |  |  |  |
| **Vaccination Status** |  |  |  |
| Vaccinated | 124(27.1) | 284(62.0) | 3.3(0.065) |
| Unvaccinated | 9(2.0) | 41(9.0) |  |
| Unknown= 237 |  |  |  |
| **Type of vaccine** |  |  |  |
| Pfizer | 38(10.0) | 148(39.1) | 80.1(<0.0001)* |
| AstraZeneca | 78(20.6) | 54(14.3) |  |
| Mixture | 2(0.5) | 59(15.6) |  |
| Unknown=316 |  |  |  |
| **Vaccine Dose** |  |  |  |
| Post first | 86(22.1) | 51(13.1) | 111.7(<0.0001)* |
| Post second | 31(8.0) | 130(33.3) |  |
| Post booster | 2(0.5) | 90(23.1) |  |
| Unknown=305 |  |  |  |
| **Hospitalization Duration** |  |  |  |
| None | 128(18.7) | 419(61.3) | 9.2(0.01)* |
| Short (≤20 days) | 25(3.65) | 40(5.85) |  |
| Long (>20 days) | 24(3.51) | 48(7.02) |  |
| Unknown=11 |  |  |  |
| **Organ Transplant patient** |  |  |  |
| Yes | 15(2.2) | 33(4.9) | 0.79(0.37) |
| No | 160(23.6) | 470(69.3) |  |
| Unknown=17 |  |  |  |
| **Ct Range** |  |  |  |
| high Ct >30 | 14(2.15) | 59(9.05) | 18.5(<0.0001)* |
| Low Ct <20 | 68(10.43) | 108(16.56) |  |
| Moderate Ct 20-30 | 91(13.96) | 312(47.85) |  |
| Unknown=43 |  |  |  |

Abbreviations: ICU, intensive care unit, Ct, cycle threshold. *Significant P value, P<0.05.

Table 9. Association of the nucleocapsid protein amino acid mutation R203M with patient demographic and clinical characteristics

| **Characteristic** | **No.(%)** | | **χ^2^ or T** |
| --- | --- | --- | --- |
|  | **R203M** **Mutation** | **Wild Type** | **(P-value)** |
| **Age (mean, SD), years** | 44.1(16.2) | 37.2(18.9) | 4.7(<0.0001)* |
| **Variant** |  |  |  |
| Alpha | 0(0) | 24(3.45) | 674.5(<0.0001)* |
| Beta | 0(0) | 23(3.31) |  |
| Delta | 175(25.18) | 4(0.58) |  |
| Delta Plus | 3(0.43) | 0(0) |  |
| Eta | 0(0) | 2(0.29) |  |
| Kappa | 1(0.14) | 0(0) |  |
| Omicron | 0(0) | 1(0.14) |  |
| Omicron BA.1 | 0(0) | 414(59.57) |  |
| Omicron BA.2 | 0(0) | 24(3.45) |  |
| Other | 0(0) | 24(3.45) |  |
| **Wave** |  |  |  |
| Delta | 150(21.6) | 442(63.6) | 293(<0.0001)* |
| Omicron | 29(4.2) | 74(10.7) |  |
| **Sex** |  |  |  |
| Male | 96(13.8) | 227(32.7) | 5.0(0.03)* |
| Female | 83(11.9) | 289(41.6) |  |
| **Nationality** |  |  |  |
| Saudi | 97(15.3) | 362(57.0) | 26.9(<0.0001)* |
| Non-Saudi | 73(11.5) | 103(16.2) |  |
| Unknown=60 |  |  |  |
| **Smoking status** |  |  |  |
| Yes | 9(1.4) | 39(6.1) | 1.4(0.22) |
| No | 159(24.7) | 437(67.9) |  |
| Unknown=51 |  |  |  |
| **Patient Status** |  |  |  |
| Deceased | 12(1.74) | 28(4.06) | 9.8(0.02)* |
| Recovered | 132(19.16) | 424(61.54) |  |
| Hospitalized | 1(0.15) | 5(0.73) |  |
| Released | 34(4.93) | 53(7.69) |  |
| Unknown=6  Unknown= |  |  |  |
| **Immunocompromised** |  |  |  |
| Yes | 37(5.5) | 114(17.0) | 0.19 (0.67) |
| No | 136(20.3) | 382(57.1) |  |
| Unknown=26 |  |  |  |
| **ICU Admission** |  |  |  |
| Yes | 34(8.6) | 58(5.0) | 7.1(0.0079)* |
| No | 140(20.7) | 445(65.7) |  |
| Unknown=18 |  |  |  |
| **Comorbidity** |  |  |  |
| Yes | 70(10.5) | 227(33.9) | 1.4(0.23) |
| No | 103(15.4) | 270(40.3) |  |
| Unknown=25 |  |  |  |
| **Diabetes mellitus** |  |  |  |
| Yes | 37(5.5) | 67(10.0) | 6.3(0.012)* |
| No | 136(21.2) | 433(64.3) |  |
| Unknown=22 |  |  |  |
| **Hypertension** |  |  |  |
| Yes | 52(7.7) | 115(17.1) | 3.4(0.064) |
| No | 121(18.0) | 385(57.2) |  |
| Unknown=22 |  |  |  |
| **Symptoms** |  |  |  |
| Asymptomatic | 2(0.3) | 30(5.0) | 7.6(0.0057)* |
| Symptomatic | 163(27.1) | 406(67.6) |  |
| Unknown= 94 |  |  |  |
| **Disease Severity** |  |  |  |
| Mild | 132(20.82) | 397(62.62) | 13.1(0.0014)* |
| Stage C | 22(3.47) | 46(7.26) |  |
| Stage D | 19(3) | 18(2.84) |  |
| Unknown=61 |  |  |  |
| **Vaccination Status** |  |  |  |
| Vaccinated | 121(26.4) | 287(62.7) | 2.0(0.15) |
| Unvaccinated | 10(2.2) | 40(8.7) |  |
| Unknown= 237 |  |  |  |
| **Type of vaccine** |  |  |  |
| Pfizer | 37(9.8) | 149(39.3) | 77.1(<0.0001)* |
| AstraZeneca | 76(20.1) | 56(14.8) |  |
| Mixture | 2(0.5) | 59(15.6) |  |
| Unknown=316 |  |  |  |
| **Vaccine Dose** |  |  |  |
| Post first | 83(21.28) | 54(13.85) | 104.3(<0.0001)* |
| Post second | 31(7.95) | 130(33.33) |  |
| Post booster | 2(0.51) | 90(23.08) |  |
| Unknown=305 |  |  |  |
| **Hospitalization Duration** |  |  |  |
| None | 126(18.4) | 421(61.6) | 9.8(0.007)* |
| Short (≤20 days) | 25(3.7) | 40(5.9) |  |
| Long (>20 days) | 24(3.5) | 48(7.0) |  |
| Unknown=11 |  |  |  |
| **Organ Transplant patient** |  |  |  |
| Yes | 15(4.9) | 33(2.2) | 1.2(0.28) |
| No | 158(23.3) | 472(69.6) |  |
| Unknown=17 |  |  |  |
| **Ct Range** |  |  |  |
| high Ct >30 | 58(8.9) | 15(2.3) | 15.9(0.0003)* |
| Low Ct <20 | 110(16.9) | 66(10.12) |  |
| Moderate Ct 20-30 | 313(48.0) | 90(13.8) |  |
| Unknown=43 |  |  |  |

Abbreviations: ICU, intensive care unit, Ct, cycle threshold. *Significant P value, P<0.05.

Table 10. Association of the nucleocapsid protein amino acid mutation D377Y with patient demographic and clinical characteristics

| **Characteristic** | **No.(%)** | | **χ^2^ or T** |
| --- | --- | --- | --- |
|  | **D377Y Mutation** | **Wild Type** | **(P-value)** |
| **Age (mean, SD), years** | 44.0(16.2) | 37.2(18.9) |  |
| **Variant** |  |  |  |
| Alpha | 0(0.0) | 24(3.5) | 664(<0.0001)* |
| Beta | 0(0.0) | 23(3.3) |  |
| Delta | 173(24.9) | 6(0.9) |  |
| Delta Plus | 3(0.4) | 0(0.0) |  |
| Eta | 0(0.0) | 2(0.3) |  |
| Kappa | 1(0.1) | 0(0.0) |  |
| Omicron | 0(0.0) | 1(0.1) |  |
| Omicron BA.1 | 0(0.0) | 414(59.6) |  |
| Omicron BA.2 | 0(0.0) | 24(3.5) |  |
| Other | 0(0.0) | 24(3.5) |  |
| **Wave** |  |  |  |
| Delta | 149(10.8) | 75(10.8) | 293.4(<0.001)* |
| Omicron | 28(4.0) | 443(63.7) |  |
| **Sex** |  |  |  |
| Male | 95(13.7) | 228(32.8) | 4.2(0.026)* |
| Female | 82(11.8) | 290 (41.7) |  |
| **Nationality** |  |  |  |
| Saudi | 97(15.3) | 362(57.0) | 24.1 (<0.0001) |
| Non-Saudi | 71(11.2) | 105(16.5) |  |
| Unknown=60 |  |  |  |
| **Smoking status** |  |  |  |
| Yes | 8(1.2) | 40(6.2) | 2.2(0.133) |
| No | 158(24.5) | 438(68.0) |  |
| Unknown=51 |  |  |  |
| **Patient Status** |  |  |  |
| Deceased | 12(1.74) | 28(4.06) | 10.4(0.0157)* |
| Recovered | 130(18.87) | 426(61.83) |  |
| Hospitalized | 1(0.15) | 5(0.73) |  |
| Released | 34(4.93) | 53(7.69) |  |
| Unknown=6  Unknown= |  |  |  |
| **Immunocompromised** |  |  |  |
| Yes | 134(20.0) | 114(17.0) | 0.11(0.73) |
| No | 106(23.9) | 384(57.4) |  |
| Unknown=26 |  |  |  |
| **ICU Admission** |  |  |  |
| Yes | 34(5.0) | 58(8.6) | 7.5(0.006)* |
| No | 138(20.4) | 447(66.0) |  |
| Unknown=18 |  |  |  |
| **Comorbidity** |  |  |  |
| Yes | 69(10.3) | 228(34.0) | 1.5(0.23) |
| No | 102(15.2) | 271(40.5) |  |
| Unknown=25 |  |  |  |
| **Diabetes mellitus** |  |  |  |
| Yes | 36(5.4) | 68(10.1) | 5.5 (0.02)* |
| No | 135(20.1) | 434(64.5) |  |
| Unknown=22 |  |  |  |
| **Hypertension** |  |  |  |
| Yes | 50(7.4) | 117(17.4) | 2.4(0.12) |
| No | 121(18.0) | 385(57.2) |  |
| Unknown=22 |  |  |  |
| **Symptoms** |  |  |  |
| Asymptomatic | 2(0.3) | 30(5.0) | 7.5(0.0006)* |
| Symptomatic | 162(27.0) | 407(67.7) |  |
| Unknown= 94 |  |  |  |
| **Disease Severity** |  |  |  |
| Mild | 130(20.5) | 399(62.93) | 13.7(0.0011)* |
| Stage C | 22(3.47) | 46(7.26) |  |
| Stage D | 19(3) | 18(2.84) |  |
| Unknown=61 |  |  |  |
| **Vaccination Status** |  |  |  |
| Vaccinated | 120(28.2) | 288(62.9) | 2.9(0.09) |
| Unvaccinated | 9(2.0) | 41(9.0) |  |
| Unknown= 237 |  |  |  |
| **Type of vaccine** |  |  |  |
| Pfizer | 38(10.0) | 148(39.1) | 73.6(<0.0001)* |
| AstraZeneca | 75(19.8) | 57(15.0) |  |
| Mixture | 2(0.5) | 59(15.6) |  |
| Unknown=316 |  |  |  |
| **Vaccine Dose** |  |  |  |
| Post first | 84(21.54) | 53(13.59) | 108.3(<0.0001)* |
| Post second | 30(7.69) | 131(33.59) |  |
| Post booster | 2(0.51) | 90(23.08) |  |
| Unknown=305 |  |  |  |
| **Hospitalization Duration** |  |  |  |
| None | 124(18.13) | 423(61.84) | 10.4(0.005)* |
| Short (≤20 days) | 25(3.65) | 40(5.85) |  |
| Long (>20 days) | 24(3.51) | 48(7.02) |  |
| Unknown=11 |  |  |  |
| **Organ Transplant patient** |  |  |  |
| Yes | 14(5.0) | 34(5.0) | 0.65(0.42) |
| No | 157(23.2) | 473(69.8) |  |
| Unknown=17 |  |  |  |
| **Ct Range** |  |  |  |
| high Ct >30 | 14(2.15) | 59(9.05) | 15.5(0.004)* |
| Low Ct <20 | 65(9.97) | 111(17.02) |  |
| Moderate Ct 20-30 | 90(13.8) | 313(48.01) |  |
| Unknown=43 |  |  |  |

Abbreviations: ICU, intensive care unit, Ct, cycle threshold. *Significant P value, P<0.05.

Table 11. Association of the nucleocapsid protein amino acid mutation G215C with patient demographic and clinical characteristics

| **Characteristic** | No.**(%)** | | **χ^2^ or T** |
| --- | --- | --- | --- |
|  | **G215C** **Mutation** | **Wild Type** | **(P-value)** |
| **Age (mean, SD), years** | 44.7(16.6) | 37.6(18.6) | 4.3(<0.0001)* |
| **Variant** |  |  |  |
| Alpha | 0(0) | 24(3.5) | 456.6(<0.0001)* |
| Beta | 0(0) | 23(3.3) |  |
| Delta | 128(18.42) | 51(7.3) |  |
| Delta Plus | 3(0.4) | 0() |  |
| Eta | 0(0) | 2(0.3) |  |
| Kappa | 0(0) | 1(0.1) |  |
| Omicron | 0(0) | 1(0.1) |  |
| Omicron BA.1 | 0(0) | 414(59.5) |  |
| Omicron BA.2 | 0(0) | 24(3.5) |  |
| Other | 0(0) | 24(3.5) |  |
| **Wave** |  |  |  |
| Delta | 107(15.4) | 117(16.8) | 180.7(<0.0001)* |
| Omicron | 24(3.4) | 447(64.3) |  |
| **Sex** |  |  |  |
| Male | 80(11.5) | 243(35.0) | 13.8(0.0002)* |
| Female | 51(7.3) | 321(46.2) |  |
| **Nationality** |  |  |  |
| Saudi | 64 | 395(32.8) | 32.9(<0.0001)* |
| Non-Saudi | 60 | 116 |  |
| Unknown=60 |  |  |  |
| **Smoking status** |  |  |  |
| Yes | 9(1.4) | 39(6.1) | 0.004(0.94) |
| No | 114(17.7) | 482(74.8) |  |
| Unknown=51 |  |  |  |
| **Patient Status** |  |  |  |
| Deceased | 8(8) | 32(4.64) | 10.7(0.0135)* |
| Recovered | 96(96) | 460(66.76) |  |
| Hospitalized | 0(0) | 6(0.87) |  |
| Released | 27(27) | 60(8.71) |  |
| Unknown=6  Unknown= |  |  |  |
| **Immunocompromised** |  |  |  |
| Yes | 27(4.0) | 124(18.5) | 0.19(0.65) |
| No | 101(15.1) | 417(62.3) |  |
| Unknown=26 |  |  |  |
| **ICU Admission** |  |  |  |
| Yes | 24(3.6) | 68(10.0) | 3.6(0.058) |
| No | 104(15.4) | 481(71.1) |  |
| Unknown=18 |  |  |  |
| **Comorbidity** |  |  |  |
| Yes | 51(7.6) | 246(37.7) | 1.3(0.25) |
| No | 77(11.5) | 296(44.2) |  |
| Unknown=25 |  |  |  |
| **Diabetes mellitus** |  |  |  |
| Yes | 24(3.6) | 80(11.9) | 1.3(0.25) |
| No | 104(15.5) | 465(69.1) |  |
| Unknown=22 |  |  |  |
| **Hypertension** |  |  |  |
| Yes | 37(5.5) | 130(19.3) | 1.4(0.23) |
| No | 91(13.5) | 415(61.7) |  |
| Unknown=22 |  |  |  |
| **Symptoms** |  |  |  |
| Asymptomatic | 2(0.3) | 30(5.0) | 4.3(0.039)* |
| Symptomatic | 122(20.3) | 447(74.4) |  |
| Unknown= 94 |  |  |  |
| **Disease Severity** |  |  |  |
| Mild | 99(15.62) | 430(67.82) | 7.4(0.0249)* |
| Stage C | 18(2.84) | 50(7.89) |  |
| Stage D | 13(2.05) | 24(3.79) |  |
| Unknown=61 |  |  |  |
| **Vaccination Status** |  |  |  |
| Vaccinated | 91(19.9) | 317(69.2) | 5.5(0.0186)* |
| Unvaccinated | 4(0.9) | 46(10.0) |  |
| Unknown= 237 |  |  |  |
| **Type of vaccine** |  |  |  |
| Pfizer | 29(7.7) | 157(41.4) | 45.5(<0.0001)* |
| AstraZeneca | 55(14.5) | 77(20.3) |  |
| Mixture | 2(0.5) | 59(15.6) |  |
| Unknown=316 |  |  |  |
| **Vaccine Dose** |  |  |  |
| Post first | 68(17.44) | 69(17.69) | 89.3(<0.0001)* |
| Post second | 19(4.87) | 142(36.41) |  |
| Post booster | 2(0.51) | 90(23.08) |  |
| Unknown=305 |  |  |  |
| **Hospitalization Duration** |  |  |  |
| None | 93(13.6) | 454(66.4) | 7.8(0.02)* |
| Short (≤20 days) | 20(2.9) | 45(6.6) |  |
| Long (>20 days) | 16(2.3) | 56(8.2) |  |
| Unknown=11 |  |  |  |
| **Organ Transplant patient** |  |  |  |
| Yes | 10(1.5) | 38 (5.6) | 0.13(0.72) |
| No | 118(17.4) | 512(75.5) |  |
| Unknown=17 |  |  |  |
| **Ct Range** |  |  |  |
| high Ct >30 | 12(1.84) | 61(9.36) | 14.4(0.0007)* |
| Low Ct <20 | 50(7.67) | 126(19.33) |  |
| Moderate Ct 20-30 | 61(9.36) | 342(52.45) |  |
| Unknown=43 |  |  |  |

Abbreviations: ICU, intensive care unit, Ct, cycle threshold. *Significant P value, P<0.05.
